# Evaluating respiratory syncytial virus immunization strategies for infants in Canada: a cost-utility analysis

**DOI:** 10.64898/2026.01.09.26343789

**Authors:** Gebremedhin B. Gebretekle, Marie Lan, Min Xi, Raphael Ximenes, Sarah A. Buchan, Elissa M Abrams, Melissa K Andrew, Nicholas Brousseau, April Killikelly, Deborah Money, Jesse Papenburg, Ellen Rafferty, Joan L Robinson, Winnie Siu, Matthew Tunis, Ashleigh R. Tuite

## Abstract

**Background:** Respiratory syncytial virus (RSV) is a leading cause of lower respiratory tract infections and hospitalizations among infants in Canada. New long-acting monoclonal antibodies (mAbs) and vaccines administered during pregnancy have expanded prevention options, yet the most cost-effective immunization program remains uncertain.

**Methods:** We updated a Canadian cost-utility model to evaluate seven seasonal RSV prevention strategies over one year (with a lifetime horizon for mortality impacts), from health system and societal perspectives. Strategies included RSVpreF vaccination in late pregnancy; targeted or universal infant mAb programs using nirsevimab or clesrovimab; and combination programs in which infants could receive protection from either RSVpreF or mAbs. Sequential incremental cost-effectiveness ratios (ICERs) were estimated in 2024 Canadian dollars per quality-adjusted life year (QALY), using a $50,000/QALY threshold. The primary analysis used immunization product list prices.

**Findings:** The most cost-effective strategy was a seasonal combination program: RSVpreF vaccination for pregnancies due during RSV season with mAb for infants at high risk (<32 weeks’ gestation), including catch-up for infants at high-risk born before the season. This strategy had an ICER of $35,408/QALY compared to seasonal mAb for infants at moderate risk (32^0/7^ to 36^6/7^ weeks’ gestation) or high-risk with catch-up. Expanding mAb to unimmunized non-high-risk infants born in-season increased the ICER to $132,131/QALY. Universal infant protection (mAb alone or combined with RSVpreF in pregnancy) was not cost-effective across analyses. RSVpreF alone was dominated. Results were most sensitive to product prices, target populations, age at administration, and RSV burden.

**Conclusions:** A seasonal combination program with RSVpreF for in-season deliveries and mAb for infants at high-risk offers the best value for money for protecting Canadian infants from RSV disease. Broader infant immunization programs may be cost-effective with substantial price reductions or in regions with higher disease burden and healthcare costs.

## 1. Introduction

Respiratory syncytial virus (RSV) is the major cause of acute lower respiratory tract infection (LRTI) and hospitalization among infants in Canada, with the highest burden among those less than six months of age (1–3). Annual RSV-associated hospitalization rates range from 5 to 28 per 1,000 in infants under 6 months of age, and 3 to 13 per 1,000 in those aged 6 to 11 months (4). Approximately 5 to 10% of hospitalized infants require intensive care (1–6). Although infants with underlying risk factors, including prematurity and chronic medical conditions have higher risks of severe disease and complications, the highest burden occurs among healthy term infants (4, 7). Seasonal surges strain pediatric healthcare capacity, leading to hospital overcrowding and a substantial economic burden (8).

Recent advances in RSV immunization have led to the introduction of long-acting monoclonal antibodies (mAbs) and a vaccine administered during pregnancy (RSVpreF) for infant protection, both of which provide infants with season-long protection from a single dose, a significant improvement over the monthly dosing required with palivizumab. In 2024, Canada’s National Advisory Committee on Immunization (NACI) recommended nirsevimab as the preferred option for infant prophylaxis, citing potential efficacy benefits and safety uncertainties regarding preterm birth with RSVpreF (9). Consequently, NACI indicated that RSVpreF may be considered on an individual basis for pregnant women and pregnant people (9). For nirsevimab, a phased rollout was recommended, prioritizing high-risk infants, with broader implementation contingent on supply, affordability, and cost-effectiveness considerations (9).

Our previous Canadian model-based economic evaluation supported NACI’s deliberations on infant RSV immunization but did not assess seasonal administration of RSVpreF or combination programs involving both RSVpreF and mAb, due to implementation uncertainties at the time (9, 10). Since then, emerging evidence supports the safety and feasibility of RSVpreF use in late pregnancy, including seasonal administration (11–16). Furthermore, the landscape of passive immunization continues to evolve, with additional long-acting mAbs (clesrovimab) demonstrating efficacy in clinical trials (17). Given these developments and ongoing implementation considerations, we performed a model-based economic evaluation of a wider range of infant RSV prevention strategies to inform policy decisions and optimize resource allocation within the Canadian healthcare system. Our objective was to identify the most cost-effective RSV prevention strategy for infants in Canada at current product list prices, and to examine how conclusions change across plausible product prices and in higher burden and higher healthcare costs settings.

## 2. Methods

### 2.1. Model Overview

We updated a previously published Canadian cost-utility model to evaluate the cost-effectiveness of RSV immunization strategies, including seasonal vaccination with RSVpreF during late pregnancy, infant mAb prophylaxis with nirsevimab or clesrovimab, and combination programs using both RSVpreF and mAb (9, 10). Key model updates included: (i) the addition of new strategies, such as seasonal RSVpreF alone and in combination with mAb, (ii) the removal of strategies dominated in the previous analysis or deemed not policy relevant (e.g., year-round programs), and (iii) the revision of key input parameters using recent Canadian data on product prices, RSV epidemiology, and updated assumptions for effectiveness and waning. The results of this analysis were used to inform forthcoming updated recommendations by NACI on RSV infant immunization programs.

A brief overview of the model structure and assumptions is provided below, with additional details available in the Supplementary Materials. A static Markov cohort model was used to estimate costs (in 2024 Canadian dollars), quality-adjusted life years (QALYs) and incremental cost-effectiveness ratios (ICERs) from both the health system and societal perspectives. The model simulated 12 monthly birth cohorts over their first year of life to capture acute RSV outcomes during their first RSV season. A lifetime horizon was applied to account for long-term mortality impacts associated with RSV, with costs and QALYs discounted at 1.5% annually, consistent with Canadian economic evaluation guidelines (18, 19).

The model structure, health states, and RSV-related outcomes followed our prior analysis (Figure 1) (10). Infants were stratified into three risk groups for RSV disease based on their week of gestational age (wGA) at birth: (i) high-risk (born <32 wGA), (ii) moderate-risk (32^0/7^ to 36^6/7^ wGA), and (iii) low-risk (≥37^0^ wGA). Our definition of high- and moderate-risk was limited to prematurity, and therefore differs from NACI’s 2024 recommendation, which prioritizes all infants born <37 wGA as well as term infants with certain chronic medical condition (e.g., chronic lung or cardiac disease) to receive mAb (9). Each infant could experience up to one medically attended lower respiratory tract infection (MA-LRTI) during their first year of life. Following MA-LRTI, infants could require outpatient care (primary care or emergency department), hospitalization (general ward or intensive care unit [ICU]), or die from RSV-related complications. RSV-attributable deaths were assumed to occur only among hospitalized infants. Immunization reduced risk of RSV-related outcomes according to product-specific effectiveness and time since immunization.

**Figure 1.**
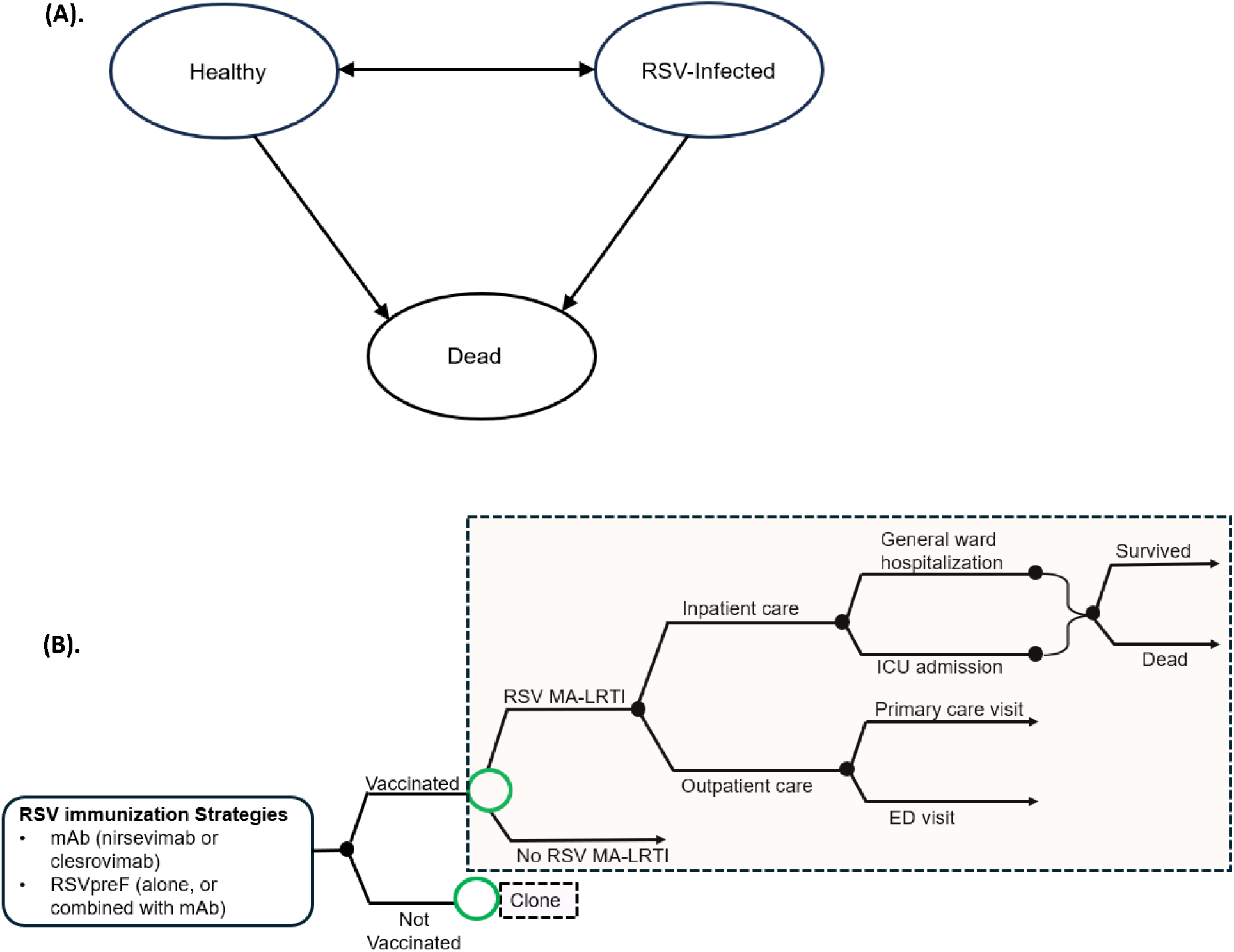
Overview of (A) health states and (B) structure of the model for RSV-related outcomes. ED: emergency department; ICU: intensive care unit; MA-LRTI: medically attended lower respiratory tract infection; mAb: monoclonal antibody; RSV: respiratory syncytial virus

### 2.2. Immunization strategies

Seven RSV immunization strategies were evaluated, all modelled as seasonal programs providing protection during the RSV season (November to April). In-season protection refers to RSVpreF vaccination for in-season due dates during the RSV season (administered at 32^0/7^ to 36^6/7^ weeks gestation) or infant mAb administered at birth during the RSV season. Catch-up protection refers to mAb administered prior to the onset of the RSV season for infants born out of season. Combination strategies incorporate RSVpreF vaccination for in-season deliveries, with mAb offered only to infants from unvaccinated pregnancies (i.e., no infant could receive both products in any currently modelled strategy). Strategy scope ranged from risk-based to broad and universal coverage. Risk-based eligibility varied by strategy: the comparator strategy included both moderate and high-risk infants (< 37 weeks’ gestation), consistent with NACI recommendations at the time of the analysis (9), whereas other risk-based programs evaluated included high-risk infants only (with the preterm eligibility limited to <32 weeks’ gestation). Broad programs included all in-season births with catch-up for high-risk infants only, while universal programs included all infants born in season and a catch-up for those born out of season.

The seven strategies were: risk-based mAb (comparator), broad mAb, universal mAb, RSVpreF only, risk-based combination, broad combination, and universal combination. Descriptions of each strategy, including eligibility, timing of protection, and product use, are provided in Table 1.

**Table 1.** Immunization strategies against RSV disease in infants

| Strategy name | Strategy details | Product | In-season protection (pregnancy or birth dose) |  |  | Off-season protection (catch-up dose)* |  |  |
| --- | --- | --- | --- | --- | --- | --- | --- | --- |
|  |  |  | HR | MR | LR | HR | MR | LR |
| Comparator |  |  |  |  |  |  |  |  |
| 1.Risk-based mAb | Seasonal mAb for infants at moderate or high-risk, with catch-up for those at moderate or high-risk | RSVpreF | NA | NA | NA | NA | NA | NA |
|  |  | mAb | ✓ | ✓ | NA | ✓ | ✓ | NA |
| Monoclonal antibodies (mAb, nirsevimab or clesrovimab) |  |  |  |  |  |  |  |  |
| 2.Broad mAb | Seasonal mAb for all infants, with catch-up for infants at high-risk only | RSVpreF | NA | NA | NA | NA | NA | NA |
|  |  | mAb | ✓ | ✓ | ✓ | ✓ | NA | NA |
| 3.Universal mAb | Seasonal mAb for all infants, with catch-up for all infants (i.e., a universal mAb program) | RSVpreF | NA | NA | NA | NA | NA | NA |
|  |  | mAb | ✓ | ✓ | ✓ | ✓ | ✓ | ✓ |
| RSVpreF vaccine |  |  |  |  |  |  |  |  |
| 4.RSVpreF only | Seasonal RSVpreF given at 32 to 36 weeks of pregnancy for those with in-season due dates | RSVpreF | NA | ✓ | ✓ | NA | NA | NA |
|  |  | mAb | NA | NA | NA | NA | NA | NA |
| Combined program of RSVpreF plus mAb |  |  |  |  |  |  |  |  |
| 5.Risk-based combination | Seasonal RSVpreF plus mAb for infants at high-risk, with catch-up for high-risk | RSVpreF | NA | ✓ | ✓ | NA | NA | NA |
|  |  | mAb | ✓ | NA | NA | ✓ | NA | NA |
| 6.Broad combination | Seasonal RSVpreF plus mAb for infants born to unvaccinated pregnant women and pregnant people, with catch-up for high-risk ** | RSVpreF | NA | ✓ | ✓ | NA | NA | NA |
|  |  | mAb | ✓ | ✓** | ✓** | ✓ | NA | NA |
| 7.Universal combination | Seasonal RSVpreF plus mAb for all infants born to unvaccinated pregnant women and pregnant people, with catch-up for all infants** | RSVpreF | NA | ✓ | ✓ | NA | NA | NA |
|  |  | mAb | ✓ | ✓** | ✓** | ✓ | ✓ | ✓ |

**HR:** high-risk (preterm infants born <32 weeks' gestational age); **LR:** low-risk (full-term infants born ≥37<sup>0</sup> weeks' gestational age); **MR:** moderate-risk (preterm infants born 32<sup>0/7</sup> to 36<sup>6/7</sup> weeks of gestational age); **NA:** not applicable; **mAb:** monoclonal antibodies
**Note:** All RSVpreF programs are seasonally administered between 32<sup>0/7</sup> and 36<sup>6/7</sup> weeks of pregnancy to pregnancies with in-season due dates (November to April) i.e., all infants born outside of the RSV season and high-risk infants (born before 32 weeks of gestational age) born during the RSV season were not protected by the RSVpreF pregnancy vaccine.
\* Catch-up protection refers to administration of monoclonal antibodies (nirsevimab or clesrovimab) to infants born outside the RSV season, given prior to the beginning of their first RSV season.
\*\* Infant protection comes from either RSVpreF or mAbs, but not from both simultaneously. RSVpreF uptake was assumed to be 65%. For infants not protected by RSVpreF (35%), mAb uptake was assumed to be 71%.

We did not model a “no immunization” strategy because the risk-based mAb strategy was considered to represent the current standard of care at the time of the analysis.

Coverage assumptions were 65% for RSVpreF, 80% for infants at high-risk receiving mAb, and 71% among those at moderate- or low-risk (20, 21).

### 2.3. Model parameters

Model inputs included parameters describing RSV epidemiology (Table 2), immunization product characteristics (Table 3), costs (S1 Table) and QALY losses (S2 Table). Data were extracted from published literature and Canadian sources, where available, supplemented by assumption when data were unavailable. Unless specified below, parameter values were consistent with those used in the previous analysis.

**Table 2.**
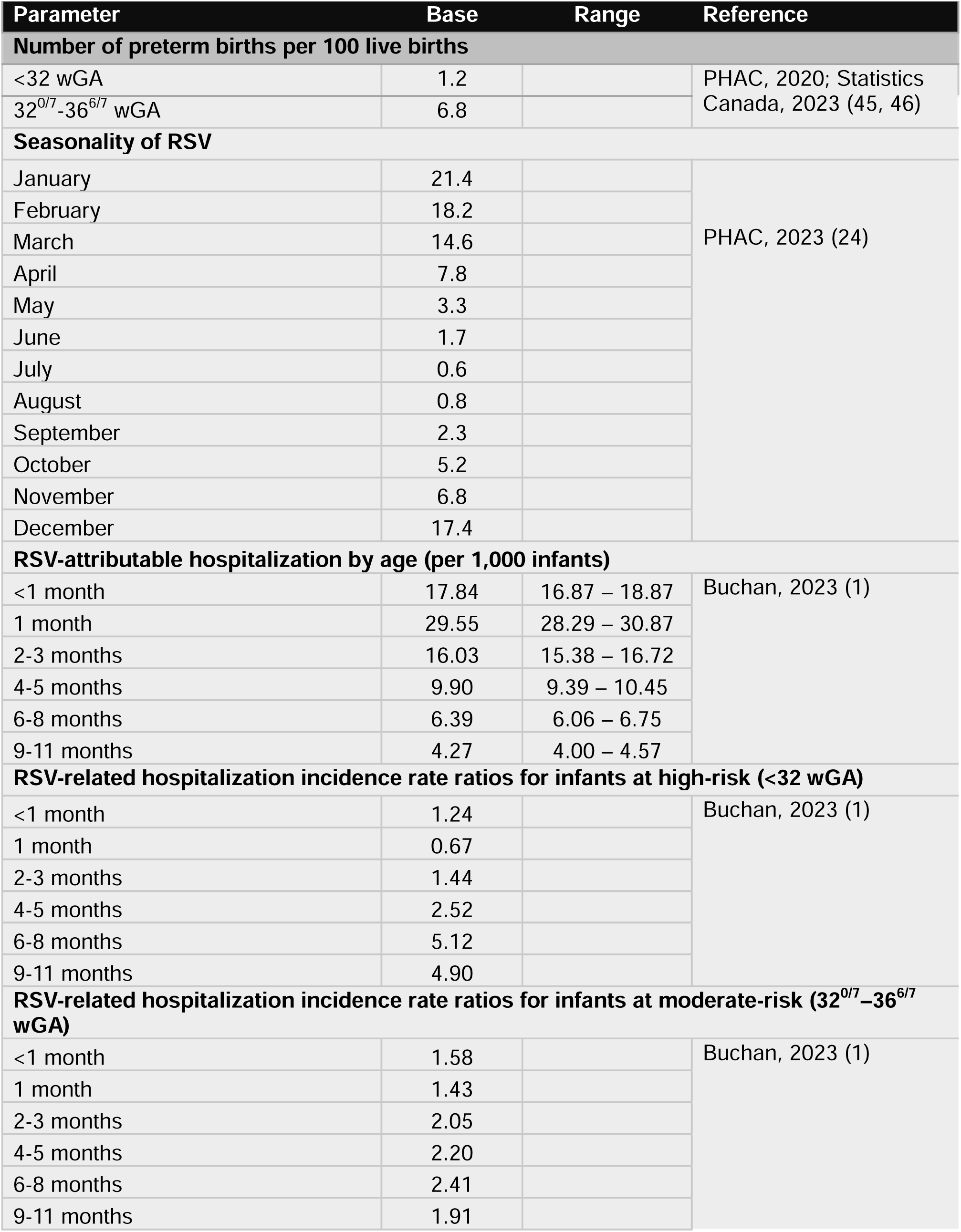
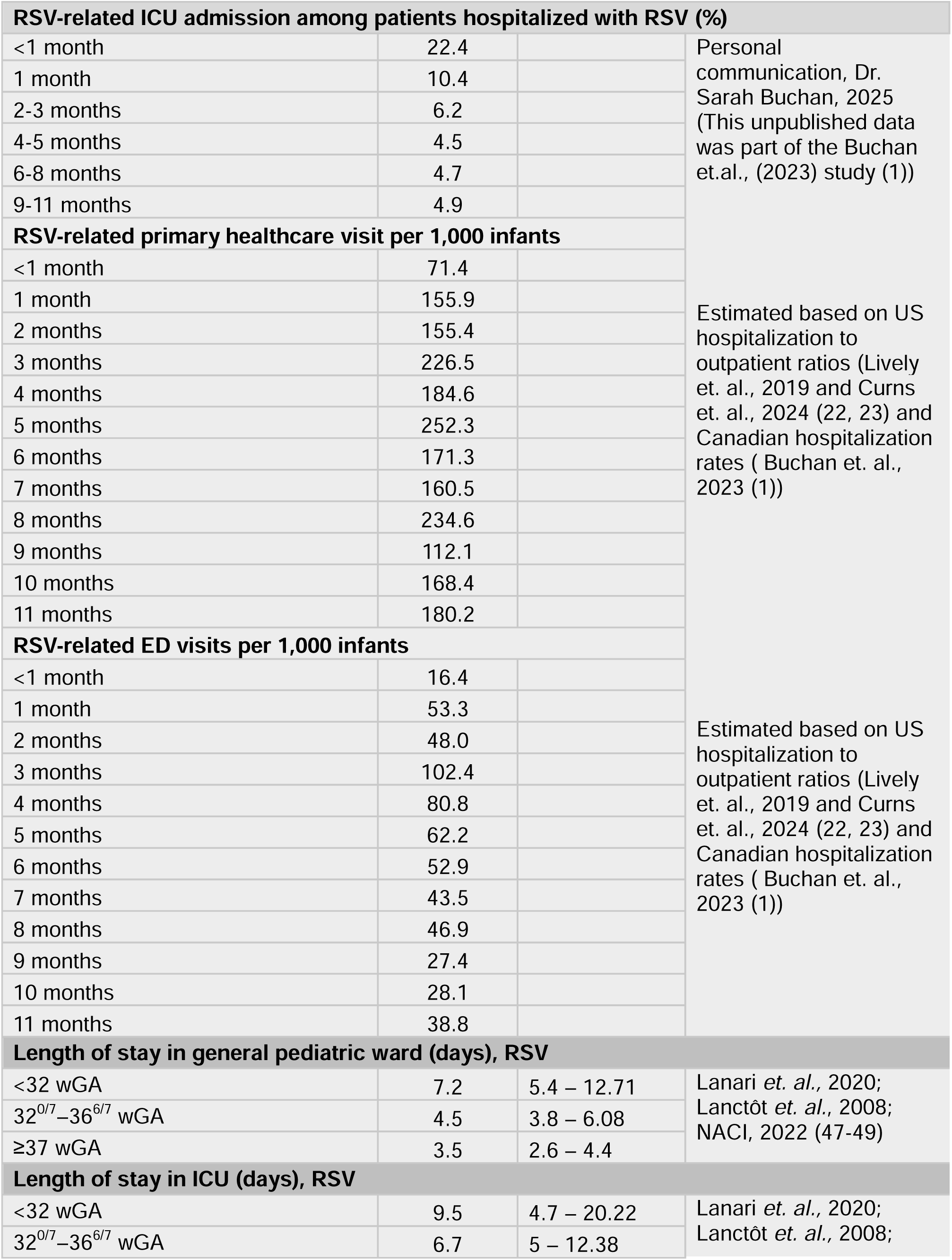
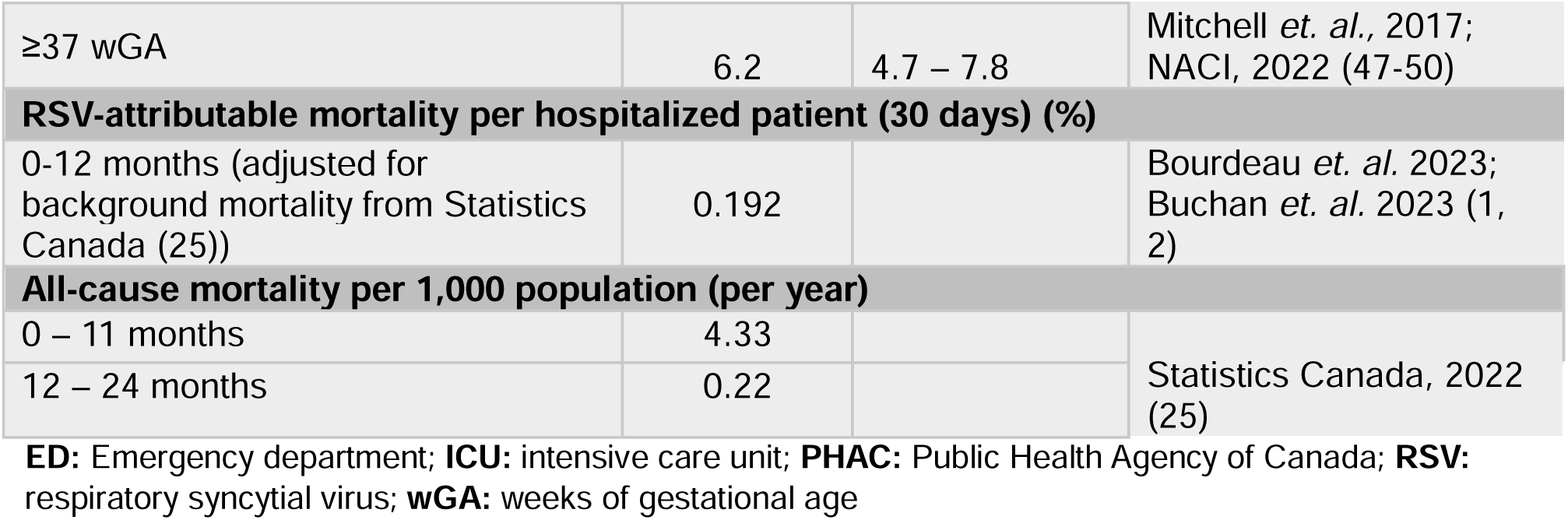
Epidemiological parameters

**Table 3.**
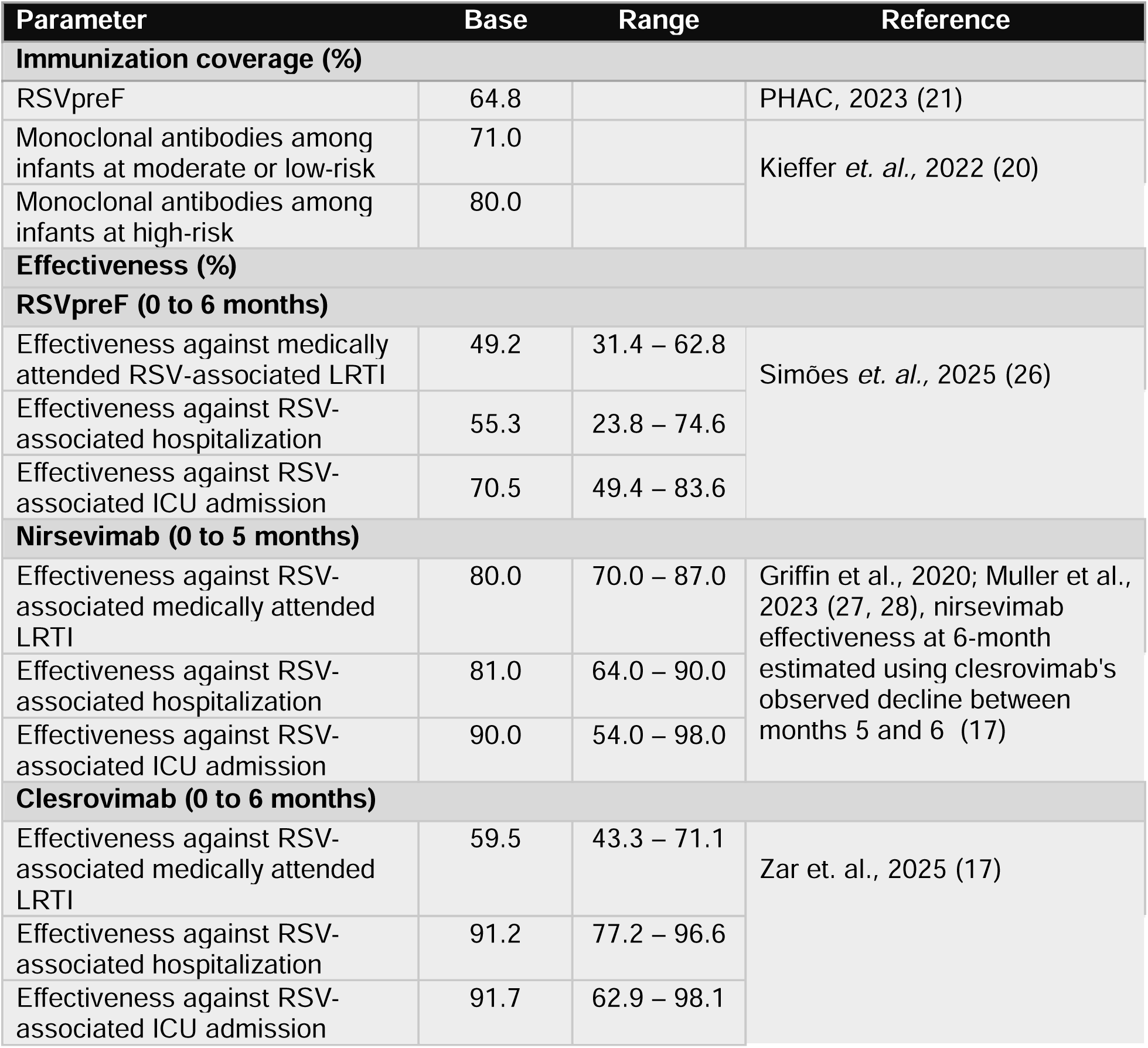
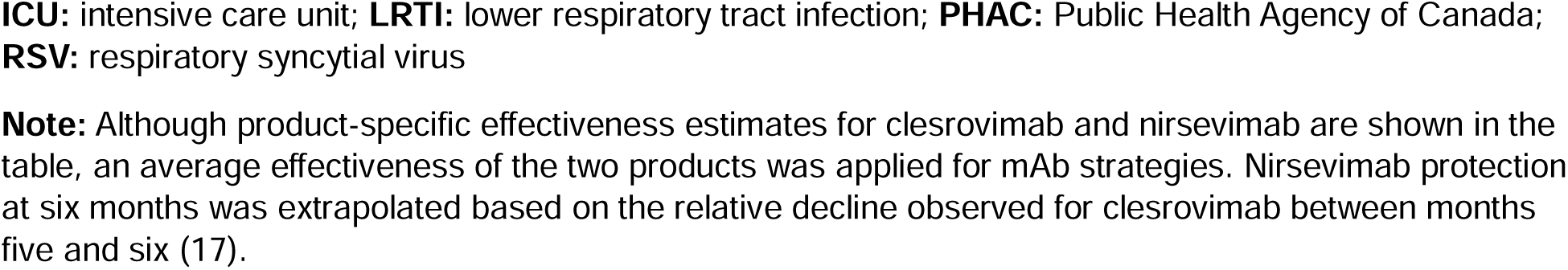
Immunization product characteristics

| Parameter | Base | Range | Reference |
| --- | --- | --- | --- |
| Immunization coverage (%) |  |  |  |
| RSVpreF | 64.8 |  | PHAC, 2023 (21) |
| Monoclonal antibodies among infants at moderate or low-risk | 71.0 |  | Kieffer *et. al.*, 2022 (20) |
| Monoclonal antibodies among infants at high-risk | 80.0 |  |  |
| Effectiveness (%) |  |  |  |
| RSVpreF (0 to 6 months) |  |  |  |
| Effectiveness against medically attended RSV-associated LRTI | 49.2 | 31.4 – 62.8 | Simões *et. al.*, 2025 (26) |
| Effectiveness against RSV-associated hospitalization | 55.3 | 23.8 – 74.6 |  |
| Effectiveness against RSV-associated ICU admission | 70.5 | 49.4 – 83.6 |  |
| Nirsevimab (0 to 5 months) |  |  |  |
| Effectiveness against RSV-associated medically attended LRTI | 80.0 | 70.0 – 87.0 | Griffin *et al.*, 2020; Muller *et al.*, 2023 (27, 28), nirsevimab effectiveness at 6-month estimated using clesrovimab's observed decline between months 5 and 6 (17) |
| Effectiveness against RSV-associated hospitalization | 81.0 | 64.0 – 90.0 |  |
| Effectiveness against RSV-associated ICU admission | 90.0 | 54.0 – 98.0 |  |
| Clesrovimab (0 to 6 months) |  |  |  |
| Effectiveness against RSV-associated medically attended LRTI | 59.5 | 43.3 – 71.1 | Zar *et. al.*, 2025 (17) |
| Effectiveness against RSV-associated hospitalization | 91.2 | 77.2 – 96.6 |  |
| Effectiveness against RSV-associated ICU admission | 91.7 | 62.9 – 98.1 |  |

**ICU:** intensive care unit; **LRTI:** lower respiratory tract infection; **PHAC:** Public Health Agency of Canada; **RSV:** respiratory syncytial virus
**Note:** Although product-specific effectiveness estimates for clesrovimab and nirsevimab are shown in the table, an average effectiveness of the two products was applied for mAb strategies. Nirsevimab protection at six months was extrapolated based on the relative decline observed for clesrovimab between months five and six (17).

#### 2.3.1. RSV epidemiology

Monthly, age-specific RSV infection rates were updated using Canadian hospitalization data (1) and outpatient-to-inpatient rate ratios from the United States (22, 23), adjusted for seasonal variation based on Canadian annual RSV distributions(24) (Table 2 and S1 Figure). Age-specific RSV-related hospitalization (1) and ICU admission rates were derived from Canadian evidence [personal communication with the first author], indicating the highest risk of severe disease occurs in the early months of life and among preterm infants. Background mortality and case-fatality rates were obtained from published sources(1, 2, 25).

#### 2.3.2. Product effectiveness and waning

We use the term “effectiveness” to refer to product-specific protection applied at the population level. These inputs were derived from recent clinical trial efficacy estimates (Table 3). In final trial analyses, RSVpreF reduced RSV-associated medically attended lower respiratory tract infection (MA-LRTI), hospitalization and ICU admissions by 49.2%, 55.3% and 70.5%, respectively, within the first 180 days of life (26). For clesrovimab, protection during the first 180 days post-dose was 59.5% against RSV MA-LRTI, 91.2% against hospitalization and 91.7% against ICU admission(17). Based on a previous meta-analysis, nirsevimab showed 80% effectiveness against MA-LRTI, 81% against hospitalization and 90% against ICU admission within 150 days post-administration (27, 28), with six-month protection extrapolated based on the decline observed for clesrovimab between months five and six(17). For mAb programs, we applied the average effectiveness of clesrovimab and nirsevimab. Protection for all products was assumed to decline over time following a sigmoidal decay curve, reaching 0% after six months (S2-S4 Figures).

#### 2.3.3. Costs and utilities

We included costs of immunization and estimated RSV-related healthcare utilization (S1 Table). Canadian list prices were informed by manufacturer-provided information available at the time of analysis: $750 for monoclonal antibodies (based on nirsevimab, as clesrovimab list price was not available) and $230 for RSVpreF. The societal perspective additionally included productivity losses due to RSV-related deaths (estimated using the human capital approach), caregiver costs, and out-of-pocket expenses. All costs are expressed in 2024 Canadian dollars, adjusted using the Consumer Price Index (29).

Health utility decrements for infants with RSV illness and their caregivers were derived from published literature (S2 Table) (30–32).

### 2.4. Analyses

We conducted sequential cost-effectiveness analyses, excluding strategies subject to absolute or extended dominance, consistent with Canadian guidelines (18, 19). Model outcomes were estimated for a hypothetical cohort of 1,000 infants. The primary analysis compared all seven strategies against each other. We also performed a secondary analysis focused on mAb-only programs (excluding programs involving use of RSVpreF), reflecting that some jurisdictions may prefer a mAb-only program. As Canada lacks an official cost-effectiveness threshold, cost-effectiveness was assessed using a $50,000 per QALY threshold, a commonly applied reference value. Model development and analyses were conducted in TreeAge Software (TreeAge Software, Inc., Williamstown, MA). Results from the health system perspective are reported here, and additional results including those from the societal perspective are available in the supplementary materials.

Scenario and sensitivity analyses assessed the impact of key parameter uncertainties. We conducted a two-way sensitivity analysis by simultaneously varying the per dose prices of RSVpreF ($50 to $230) and mAb ($50 to $750). For each price combination, we identified the most cost-effective strategy at a cost-effectiveness threshold of $50,000 per QALY, using a net monetary benefit framework that combines incremental QALYs and costs into a single value for comparison. Additional scenarios examined (i) extended duration of protection for all products, with sigmoidal waning to 0% at 12 months and (ii) higher RSV burden (33, 34) and higher healthcare costs(35, 36), representative of some remote and isolated communities in Canada (S3 Table).

## 3. Results

In the primary analysis, the risk-based combination strategy (seasonal RSVpreF plus mAb for infants at high-risk, with catch-up for infants at high-risk born out of the RSV season), was the most cost-effective option, with an ICER of $35,408 per QALY relative to the risk-based mAb strategy (mAb for infants at moderate or high-risk) (Figure 2 and S4 Table). Expanding to the broad combination strategy, which also provides mAb for infants not at high-risk who were born during the RSV season to unvaccinated pregnant women and pregnant people, increased the ICER to $132,131 per QALY compared to the risk-based combination strategy. Universal infant protection strategies (universal mAb or universal combination) were not cost-effective at current product list prices, with ICERs far exceeding commonly used thresholds. A strategy of RSVpreF only (RSVpreF given at 32-36 weeks of pregnancy for in-season due dates) was dominated, producing fewer health gains at higher costs than other strategies. ICERs from the societal perspective were generally lower than those from the health system perspective but this did not qualitatively change the conclusions.

**Figure 2.**
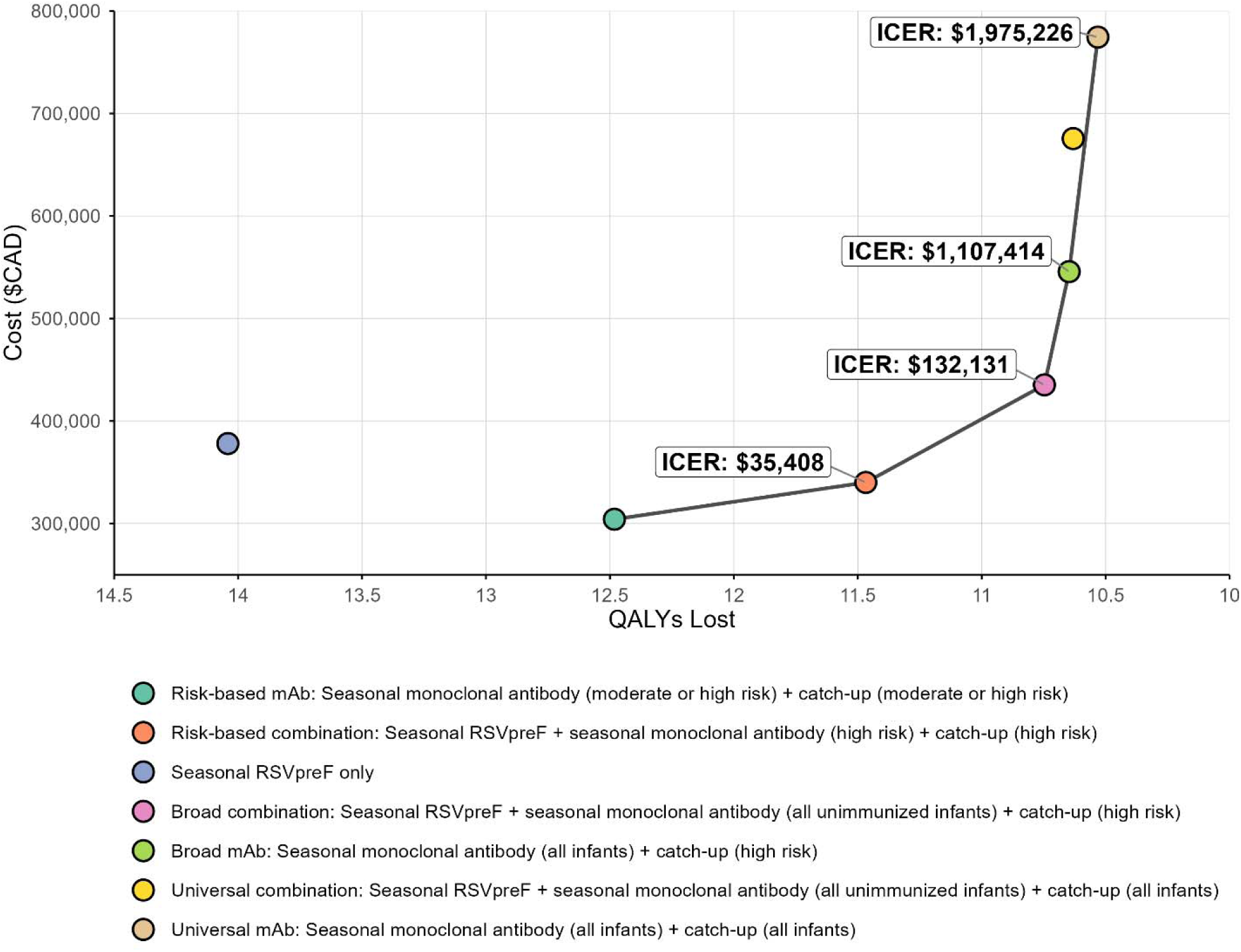
Results of the primary cost-utility analysis showing costs, QALY losses, and sequential incremental cost-effectiveness ratios, in which monoclonal antibody and/or RSVpreF programs are compared stepwise to identify the most efficient options from a health system perspective. The solid line (cost-effectiveness frontier) connects the non-dominated strategies and represents the sequence of strategies that provided the greatest incremental health gain for each additional dollar spent. *Note: All RSVpreF programs are seasonally administered between 32^0/7^ and 36^6/7^ weeks of pregnancy to those with in-season due dates (November to April) i.e., all infants born outside of the RSV season and high-risk infants (born before 32 weeks of gestational age) born during the RSV season were not protected by the RSVpreF pregnancy vaccine. Seasonal monoclonal antibody for “all unimmunized infants” refers to administration of monoclonal antibody (nirsevimab or clesrovimab) both to infants at high-risk and to those not at high-risk who were born to unvaccinated pregnant women and pregnant people during the RSV season. For combination programs, infants received protection from either product (RSVpreF or monoclonal antibody), not both*.

In the secondary analysis, which examined mAb-only strategies (i.e., excluded RSVpreF), the risk-based mAb strategy was the most cost-effective option (S5 Figure). Expanding in-season mAb to all infants, while restricting catch-up to infants at high-risk (broad mAb strategy) resulted in an ICER of $131,668 per QALY compared to the risk-based mAb strategy; to reach a cost-effectiveness threshold of $50,000/QALY a price reduction of at least 55% (to below $341 per dose) would be required. A universal mAb program covering all infants (both those born in-season and via catch-up for those born out of season), was not cost-effective, with a sequential ICER exceeding $1.9 million per QALY compared to the broad mAb strategy.

The two-way sensitivity analysis showed the risk-based combination strategy remained optimal when the mAb price exceeded $350 per dose (Figure 3). When the mAb price fell below $100 per dose, the optimal strategy shifted to broad mAb. For mAb prices between $100 and $350 per dose, the optimal strategy depended on the price of RSVpreF: at lower RSVpreF prices, a broad combination program was preferred (seasonal RSVpreF vaccination plus mAb for all unimmunized infants with catch-up for infants at high-risk); whereas at higher RSVpreF prices, a broad mAb strategy (seasonal mAb for all infants with catch-up for infants at high-risk) was more cost-effective.

**Figure 3.**
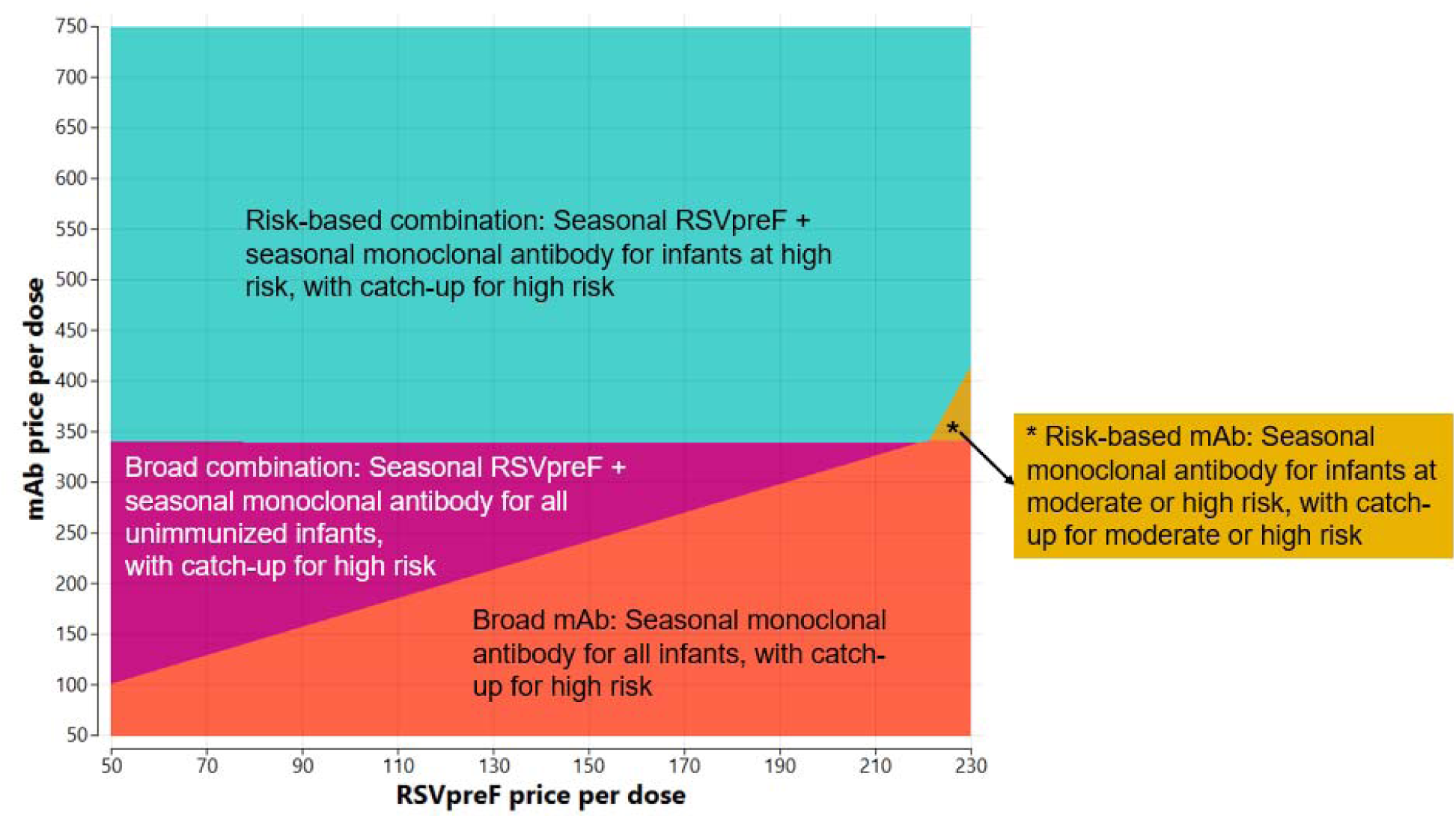
Two-way sensitivity analysis showing the impact of varying prices for monoclonal antibody and RSVpreF on the cost-effectiveness of all evaluated strategies, using a $50,000 per QALY cost-effectiveness threshold. Note: The colored areas indicate the most cost-effective strategy for a given combination of prices, showing how product pricing influences the optimal program choice. All RSVpreF programs are seasonally administered between 32^0/7^ and 36^6/7^ weeks of pregnancy to those with in-season due dates (November to April) i.e., all infants born outside of the RSV season and high-risk infants (born before 32 weeks of gestational age) born during the RSV season were not protected by the RSVpreF pregnancy vaccine. Seasonal monoclonal antibody for “all unimmunized infants” refers to administration of monoclonal antibody (nirsevimab or clesrovimab) both to infants at high-risk and to those not at high-risk who were born to unvaccinated pregnant women and pregnant people during the RSV season. For combination programs, infants received protection from either product (RSVpreF or monoclonal antibody), not both.

Assuming a longer duration of protection improved cost-effectiveness (i.e., lowered sequential ICERs) across all strategies but did not change the overall ranking of preferred strategies or conclusions regarding the optimal strategy (S6 Figure). Under this assumption, the ICER for the risk-based combination program decreased to $12,000/QALY (versus $35,408 per QALY in the primary analysis). Expanding this combination strategy to include mAb for unimmunized non-high-risk infants born in-season (broad combination strategy) resulted in an ICER of $115,101 per QALY (versus $132,131 per QALY in the primary analysis) compared to the risk-based combination strategy.

In the higher RSV burden and higher healthcare costs scenario, the broad mAb strategy (all in-season with catch-up for high-risk) was optimal at current list prices (S7 Figure). Results of the corresponding two-way sensitivity analysis are provided in the supplementary material (S8 Figure).

## 4. Discussion

This model-based economic evaluation provides updated evidence on the cost-effectiveness of infant RSV immunization strategies in Canada. At current product list prices, a combination program that uses seasonal RSVpreF during pregnancy alongside mAb for infants at high-risk of RSV, generally provided the greatest health gains per dollar spent. Specifically, we found that a risk-based combination program with seasonal RSVpreF administered at 32 to 36 weeks of gestation for pregnancies due in-season, plus mAb for infants at high-risk, including catch-up doses for high-risk infants born before the season, was the most cost-effective strategy. When strategies using RSVpreF were excluded, a risk-based mAb strategy was preferred; a broad mAb strategy (seasonal mAb for infants with catch-up for high-risk) became cost-effective only with substantial mAb price reductions (e.g., over 55% reduction), or in higher RSV burden and healthcare costs settings. RSVpreF-only strategies were never cost-effective because they leave high-risk infants unprotected.

This analysis builds on our previous evaluation of RSVpreF and nirsevimab for Canadian infants compared to the palivizumab program for infants at high-risk (10), which informed NACI’s initial guidance on infant RSV immunizations. That earlier analysis found that a risk-based nirsevimab program was the most cost-effective, whereas year-round RSVpreF or universal nirsevimab programs were not cost-effective at assumed prices. The current analysis updates and extends that work by evaluating seasonal RSVpreF and combination strategies, which have been demonstrated to be feasible based on implementation in other jurisdictions (13, 15, 16, 37). It also incorporates updated data on key inputs, including product prices, RSV burden, and emerging evidence on clesrovimab and other immunization product effectiveness and durability, to provide an assessment of cost-effectiveness for currently relevant RSV prevention strategies for infants in Canada.

Our results are consistent with recent Canadian studies favoring risk-based mAb or targeted combination strategies over universal infant programs (38–40). In Ontario, Shoukat et al. (2023) reported that a universal nirsevimab program would be cost-effective compared to no intervention only if the per dose price fell below $215 from a healthcare payer perspective (or $290 from a societal perspective) using a $50,000 per QALY threshold (40). They also demonstrated that a year-round RSVpreF combined with targeted nirsevimab for high-risk infants could achieve comparable reduction in inpatient care (pediatric ward and ICU admissions) and death as a universal nirsevimab program, but with lower budget impact (40). A pan-Canadian evaluation by Bugden et al. (2025) similarly concluded that replacing existing palivizumab programs with nirsevimab was cost-saving nationwide, but expanding coverage more broadly was highly context dependent: in southern Canada, a universal nirsevimab program required a per-dose price below $112 to be cost-effective at a $100,000 per QALY threshold, while universal coverage was cost-effective in Nunavut/Nunavik due to higher RSV burden and healthcare costs (39). In contrast to our findings, combined RSVpreF and nirsevimab programs were not cost-effective, likely because they assumed a higher RSVpreF price ($298.70 per dose) and lower nirsevimab price ($533.29 per dose) than used in our analysis (39). In British Columbia, replacing the palivizumab program with nirsevimab for moderate or high-risk infants was cost-saving (dominant) at assumed prices ($450 per dose for nirsevimab and $3,600 per RSV season for palivizumab), while an all-infant nirsevimab program required prices below $110 per dose to be cost-effective (38). Across most price combinations, either seasonal RSVpreF plus nirsevimab for high-risk infants or nirsevimab for moderate or high-risk infants were preferred strategies (38). A manufacturer-sponsored Canadian study comparing universal nirsevimab to palivizumab for high-risk infants suggested a maximum price threshold of $536 per dose at $50,000 per QALY threshold but did not consider other risk-based or combination program options, which may partly explain the higher price threshold for universal coverage in this study (41). These Canadian findings collectively show that the cost-effectiveness of infant RSV prevention strategies is influenced by several factors, including product prices, target population risk, and timing of product administration.

International studies show similar variability, with context influencing results. ICERs for strategies using nirsevimab and RSVpreF range from cost-saving to well above commonly used thresholds, reflecting differences in RSV burden, prices, comparator strategies, product pricing and other assumptions(42–44). Programs focused on higher-risk infants or higher burden regions are generally cost-effective, whereas universal infant programs often require large product price reductions to meet accepted cost-effectiveness thresholds.

Our study has some limitations. In the absence of evidence on the impact of the immunizing agents on RSV spread or indirect protection, we employed a static modeling approach that does not incorporate dynamic transmission effects. Our model focused exclusively on infant outcomes and did not assess potential direct benefits of RSVpreF vaccination for pregnant women and pregnant people. Additionally, our definition of high-risk only included key population groups included in current Canadian recommendations and did not include term infants with chronic medical conditions, who are currently recommended for prioritized prophylaxis under NACI guidelines. The model did not include certain health outcomes, including upper respiratory tract infections, or long-term complications like recurrent wheezing and asthma, due to uncertain intervention effects on these endpoints. If RSVpreF vaccine uptake during pregnancy is lower than assumed, more infants would require mAb protection in some combination strategies; higher mAb uptake would increase program costs and would be expected to reduce cost-effectiveness of these combination strategies. Finally, product list prices likely overestimate negotiated procurement prices in Canada, which are confidential. We addressed this by conducting sensitivity analyses across plausible price ranges.

## 5. Conclusions

Seasonal RSVpreF vaccine during pregnancy, combined with mAb for infants at higher risk of severe RSV disease, offers the greatest value for money among the strategies evaluated. Broader infant immunization programs could be justified in settings with high disease burden and healthcare costs, or with substantially lower product prices; however, a universal infant program is unlikely to be cost-effective at current product prices. These findings support phased implementation beginning with a targeted combination approach, with flexibility to expand program scope as affordability allows, while recognizing that jurisdictions may also consider non-economic factors such as equity, feasibility and alignment with existing program structures.

## Data Availability

All data inputs used in the model are from published sources and aggregate inputs needed to reproduce the analysis are provided.

## Acknowledgments

The authors would like to thank the NACI Secretariat team, and the NACI RSV Working Group members.

## Author contributions

**Conceptualization:** GBG, MX, RX, SAB, EMA, MKA, NB, AK, DM, JP, ER, JLR, WS, MT, ART

**Data curation:** GBG, ML

**Formal analysis / Modelling:** GBG, ART

## Manuscript drafting: **GBG**

**Manuscript review and editing:** ML, MX, RX, SAB, EMA, MKA, NB, AK, DM, JP, ER, JLR, WS, MT, ART

## Data Availability

All data inputs used in the model are from published sources and aggregate inputs needed to reproduce the analysis are provided. No individual-level health data were used.

## References

1. Buchan SA, Chung H, To T, Daneman N, Guttmann A, Kwong JC, et al. Estimating the incidence of first RSV hospitalization in children born in Ontario, Canada. J Pediatric Infect Dis Soc. 2023;12(7):421–30.

2. Bourdeau M, Vadlamudi NK, Bastien N, Embree J, Halperin SA, Jadavji T, Kazmi K, Langley JM, Lebel MH, Le Saux N, Moore D, Morris SK, Pernica JM, Robinson J, Sadarangani M, Bettinger JA, Papenburg J; Canadian Immunization Monitoring Program Active (IMPACT) Investigators. Pediatric RSV-Associated Hospitalizations Before and During the COVID-19 Pandemic. JAMA Netw Open. 2023 Oct 2;6(10):e2336863. doi: 10.1001/jamanetworkopen.2023.36863. PMID: 37792376; PMCID: PMC10551765.

3. Pisesky A, Benchimol EI, Wong CA, Hui C, Crowe M, Belair MA, et al. Incidence of Hospitalization for Respiratory Syncytial Virus Infection amongst Children in Ontario, Canada: A Population-Based Study Using Validated Health Administrative Data. PLoS One. 2016;11(3):e0150416.

4. Abrams EM, Doyon-Plourde P, Davis P, Brousseau N, Irwin A, Siu W, Killikelly A. Burden of disease of respiratory syncytial virus in infants, young children and pregnant women and people. Can Commun Dis Rep. 2024;50(1-2):1–15.

5. Buchan SA, Chung H, Karnauchow T, McNally JD, Campitelli MA, Gubbay JB, et al. Characteristics and Outcomes of Young Children Hospitalized With Laboratory-confirmed Influenza or Respiratory Syncytial Virus in Ontario, Canada, 2009-2014. Pediatr Infect Dis J. 2019;38(4):362–9.

6. Paramo MV, Ngo LPL, Abu-Raya B, Reicherz F, Xu RY, Bone JN, et al. Respiratory syncytial virus epidemiology and clinical severity before and during the COVID-19 pandemic in British Columbia, Canada: a retrospective observational study. Lancet Reg Health Am. 2023;25:100582.

7. Wingert A, Pillay J, Moore DL, Guitard S, Vandermeer B, Dyson MP, et al. Burden of illness in infants and young children hospitalized for respiratory syncytial virus: A rapid review. Can Commun Dis Rep. 2021;47(9):381–96.

8. Mitchell I, Defoy I, Grubb E. Burden of Respiratory Syncytial Virus Hospitalizations in Canada. Can Respir J. 2017;2017:4521302. doi: 10.1155/2017/4521302.

9. National Advisory Committee on Immunization. Statement on the prevention of respiratory syncytial virus (RSV) disease in infants. Ottawa (ON): Public Health Agency of Canada. 2024.

10. Gebretekle BG, Yeung MW, Ximenes R, Cernat A, Alison ES, Killikelly A, et al. Cost-effectiveness of RSVpreF vaccine and nirsevimab for the prevention of respiratory syncytial virus disease in Canadian infants. Vaccine. 2024;42(21):126164.

11. DeSilva, M., et al. (2025). Prenatal RSVpreF Vaccine Safety 2023–2024 Respiratory Season. Presented at the CDC Advisory Committee on Immunization Practices (ACIP), June 25–26, 2025. Retrieved from https://www.cdc.gov/acip/downloads/slides-2025-06-25-26/04a-DeSilva-Mat-Peds-RSV-508.pdf.

12. Son M, Riley LE, Staniczenko AP, Cron J, Yen S, Thomas C, et al. Nonadjuvanted Bivalent Respiratory Syncytial Virus Vaccination and Perinatal Outcomes. JAMA Netw Open. 2024;7(7):e2419268.

13. Haute Autorité de Santé (HAS). RSV infection vaccination recommendation for pregnant women [Internet]. Saint-Denis, France: Haute Autorité de Santé; 2024 [cited 2025 Nov 4]. Available from: https://www.has-sante.fr/jcms/p_3505344/en/rsv-infection-vaccination-recommendation-for-pregnant-women.

14. Fleming-Dutra KE, Jones JM, Roper LE, Prill MM, Ortega-Sanchez IR, Moulia DL, et al. Use of the Pfizer Respiratory Syncytial Virus Vaccine During Pregnancy for the Prevention of Respiratory Syncytial Virus-Associated Lower Respiratory Tract Disease in Infants: Recommendations of the Advisory Committee on Immunization Practices - United States, 2023. MMWR Morb Mortal Wkly Rep. 2023;72(41):1115–22.

15. UK Health Security Agency. Respiratory syncytial virus (RSV) maternal vaccination coverage in England: April 2025. GOV.UK; updated 27 November 2025. Accessed 17 December 2025. Available from: https://www.gov.uk/government/publications/rsv-maternal-vaccination-coverage-in-england/respiratory-syncytial-virus-rsv-maternal-vaccination-coverage-in-england-april-2025.

16. Trusinska D, Lee B, Ferdous S, Kwok HHY, Gordon B, Gao J, et al. Real-world uptake of nirsevimab, RSV maternal vaccine, and RSV vaccines for older adults: a systematic review and meta-analysis. EClinicalMedicine. 2025;84:103281.

17. Zar HJ, Simoes EAF, Madhi SA, Ramilo O, Senders SD, Shepard JS, et al. Clesrovimab for Prevention of RSV Disease in Healthy Infants. N Engl J Med. 2025;393(13):1292–303.

18. National Advisory Committee on Immunization. Guidelines for the economic evaluation of vaccination programs in Canada. Ottawa (ON): Public Health Agency of Canada. Available at: https://www.canada.ca/en/public-health/services/immunization/national-advisory-committee-on-immunization-naci/methods-process/incorporating-economic-evidence-federal-vaccine-recommendations/guidelines-evaluation-vaccination-programs-canada.html. 2023.

19. Canada’s Drug Agency. Guidelines for the Economic Evaluation of Health Technologies: Canada [Internet]. Ottawa: Canada’s Drug Agency; 2021 [cited 2025 Nov 4]. Available from: https://www.cda-amc.ca/guidelines-economic-evaluation-health-technologies-canada-0.

20. Kieffer A, Beuvelet M, Sardesai A, Musci R, Milev S, Roiz J, Lee JKH. Expected Impact of Universal Immunization With Nirsevimab Against RSV-Related Outcomes and Costs Among All US Infants in Their First RSV Season: A Static Model. J Infect Dis. 2022;226(Suppl 2):S282–S92.

21. Public Health Agency of Canada. Results of the Survey on Vaccination during Pregnancy 2021. Ottawa (ON); 2023. 2023.

22. Curns AT, Rha B, Lively JY, Sahni LC, Englund JA, Weinberg GA, et al. Respiratory Syncytial Virus-Associated Hospitalizations Among Children <5 Years Old: 2016 to 2020. Pediatrics. 2024;153(3).

23. Lively JY, Curns AT, Weinberg GA, Edwards KM, Staat MA, Prill MM, et al. Respiratory Syncytial Virus-Associated Outpatient Visits Among Children Younger Than 24 Months. J Pediatric Infect Dis Soc. 2019;8(3):284–6.

24. Public Health Agency of Canada (PHAC). Respiratory virus detections in Canada [Internet]. Ottawa: Public Health Agency of Canada; 2024 [cited 2025 Nov 4]. Available from: https://www.canada.ca/en/public-health/services/surveillance/respiratory-virus-detections-canada.html.

25. Statistics Canada. Life Tables, Canada, Provinces and Territories 1980/1982 to 2018/2020 (three-year estimates), and 1980 to 2020 (single-year estimates). Available at: https://www150.statcan.gc.ca/n1/pub/84-537-x/84-537-x2021001-eng.htm.

26. Simoes EAF, Pahud BA, Madhi SA, Kampmann B, Shittu E, Radley D, et al. Efficacy, Safety, and Immunogenicity of the MATISSE (Maternal Immunization Study for Safety and Efficacy) Maternal Respiratory Syncytial Virus Prefusion F Protein Vaccine Trial. Obstet Gynecol. 2025;145(2):157–67.

27. Griffin MP, Yuan Y, Takas T, Domachowske JB, Madhi SA, Manzoni P, et al. Single-Dose Nirsevimab for Prevention of RSV in Preterm Infant. N Engl J Med. 2020;383(5):415–25.

28. Muller WJ, Madhi SA, Seoane Nunez B, Baca Cots M, Bosheva M, Dagan R, et al. Nirsevimab for Prevention of RSV in Term and Late-Preterm Infants. N Engl J Med. 2023;388(16):1533–4.

29. Statistics Canada. Table 18-10-0005-01. Consumer Price Index, annual average, not seasonally adjusted [Internet]. Ottawa (ON): Government of Canada; 2023 Jan 17 [cited 2024 Jan 05]. Available at: 10.25318/1810000501-eng.

30. Glaser EL, Hariharan D, Bowser DM, et al. Impact of Respiratory Syncytial Virus on Child, Caregiver, and Family Quality of Life in the United States: Systematic Literature Review and Analysis The Journal of Infectious Diseases. 2022;226(Suppl 2):S236–S245.

31. Roy, LM. Deriving health utility weights for infants with Respiratory Syncytial Virus (RSV). University of British Columbia. Thesis/Dissertation, 2013. Accessed 20 June 2023. Available at: https://open.library.ubc.ca/collections/ubctheses/24/items/1.0074259.

32. Regnier SA. Respiratory syncytial virus immunization program for the United States: impact of performance determinants of a theoretical vaccine. Vaccine. 2013;31(40):4347–54.

33. Singleton R, Semling C, Parker J. Palivizumab Prophylaxis in Alaska for the 2022–23 RSV Season. States of Alaska Epidemiology Bulletin No. 11; 2022. Accessed 25 July 2023. Available at: https://epi.alaska.gov/bulletins/docs/b2022_11.pdf.

34. Karron R SR, Bulkow L et al. Severe respiratory syncytial virus disease in Alaska Native children. J Infect Dis. 1999;180:41–9.

35. Banerji A, Ng K, Moraes TJ, Panzov V, Robinson J, Lee BE. Cost-effectiveness of palivizumab compared to no prophylaxis in term infants residing in the Canadian Arctic. CMAJ Open. 2016;4(4):E623–E633.

36. Nourbakhsh S, Shoukat A, Zhang K, Poliquin G, Halperin D, Sheffield H, et al. Effectiveness and cost-effectiveness of RSV infant and maternal immunization programs: A case study of Nunavik, Canada. EClinicalMedicine. 2021;41:101141.

37. Jacobson KB, Watson AJ, Merchant M, Fireman B, Zerbo O, Klein NP. Uptake of Maternal RSV Vaccination and Infant Nirsevimab Among Infants Born October 2023 to March 2024. JAMA Netw Open. 2025;8(1):e2453696.

38. Taleshi J, Paramo MV, Watts A, Chilvers M, Wong JMH, Piszczek J, et al. Cost-Effectiveness of Infant and Maternal RSV Immunization Strategies, in British Columbia, Canada. Vaccine. 2025;68:127936.

39. Bugden S, Mital S, Nguyen HV. Cost-effectiveness of nirsevimab and maternal RSVpreF for preventing respiratory syncytial virus disease in infants across Canada. BMC Med. 2025;23(1):102.

40. Shoukat A, Abdollahi E, Galvani AP, Halperin SA, Langley JM, Moghadas SM. Cost-effectiveness analysis of nirsevimab and maternal RSVpreF vaccine strategies for prevention of Respiratory Syncytial Virus disease among infants in Canada: a simulation study. Lancet Reg Health Am. 2023;28:100629.

41. Shin T, Lee JK, Kieffer A, Greenberg M, Wu J. Health economic evaluation of implementing a universal immunization program with nirsevimab compared to standard of care for the prevention of respiratory syncytial virus disease in Canadian infants. Hum Vaccin Immunother. 2025;21(1):2480875.

42. Brown R, Tiggelaar S, Tsoi B, Cromwell I. Cost-effectiveness of nirsevimab for the prevention of respiratory syncytial virus outcomes in infants: CADTH Health Technology Review. Ottawa (ON): Canadian Agency for Drugs and Technologies in Health; 2023. Available from: https://www.cda-amc.ca/cost-effectiveness-nirsevimab-prevention-respiratory-syncytial-virus-outcomes-in-infants.

43. Verbrugghe S, Mahood Q, Tiggelaar S. Cost-Effectiveness of an RSVpreF Vaccine for Prevention of Respiratory Syncytial Virus Outcomes in Infants. Ottawa (ON): Canadian Agency for Drugs and Technologies in Health; 2023 Aug. Report No.: HE0044-000. Available from: https://www.cda-amc.ca/sites/default/files/hta-he/HE0044%20RSV%20Immunization%20During%20Pregnancy.pdf.

44. Zhu B, Lu Y, Zhou Y, Li W, Wu Y, Bao Y, Lu Y. Economic evaluations of RSV preventive strategies: a systematic review of cost-effectiveness and modeling approaches. Front Public Health. 2025;13:1672683.

45. Statistics Canada. Table 13-10-0425-01 Live births, by weeks of gestation. Available at: 10.25318/1310042501-eng.

46. Public Health Agency of Canada. Perinatal Health Indicators Data Tool, 2020 Edition. Centre for Surveillance and Applied Research, Public Health Infobase. Ottawa (ON): Public Health Agency of Canada, 2020. Available at: https://health-infobase.canada.ca/phi/data-tool/index?Dom=1.

47. National Advisory Committee on Immunization. Recommended use of palivizumab to reduce complications of respiratory syncytial virus infection in infants. (2022). Available at: https://www.canada.ca/en/public-health/services/publications/vaccines-immunization/palivizumab-respiratory-syncitial-virus-infection-infants.html#a5.5.

48. Lanctôt KL, Masoud ST, Paes BA, Tarride JE, et al. The cost-effectiveness of palivizumab for respiratory syncytial virus prophylaxis in premature infants with a gestational age of 32–35 weeks: a Canadian-based analysis. Curr Med Res Opin. 2008;24(11):3223–37. doi: 10.1185/03007990802484234.

49. Lanari M, Anderson EJ, Sheridan-Pereira M, Carbonell-Estrany X, et al. Burden of respiratory syncytial virus hospitalisation among infants born at 32–35 weeks’ gestational age in the Northern Hemisphere: pooled analysis of seven studies. Epidemiol Infect. 2020;148:e170. doi: 10.1017/S0950268820001661.

50. Mitchell I, Defoy I, Grubb E. Burden of Respiratory Syncytial Virus Hospitalizations in Canada. Can Respir J. 2017;2017:4521302.

