## Supplementary Materials for "Evaluating respiratory syncytial virus immunization strategies for infants in Canada: a cost-utility analysis"

This Supplementary material provides additional information regarding parameterisation of the model, and includes figures and tables supporting results reported in the main text, as well as the results of secondary analyses.

**Content**

1. Model parameter inputs and assumptions
   1.1. RSV incidence rate by age and calendar month (S1 Figure)
   1.2. Cost parameters (S1 Table)
   1.3. RSV-associated health utility decrements (S2 Table)
   1.4. Inputs for higher burden and higher medical costs scenario (S3 Table)
   1.5. Effectiveness and waning assumptions (S2-S4 Figures)
2. Results
   2.1. Primary analysis results (S4 Table)
   2.2. Secondary analysis results (mAb only programs) (S5 Figure)
   2.3. Longer protection duration scenario results (S6 Figure)
   2.4. Higher RSV burden and higher medical costs scenario results (S7-S8 Figures)
   2.5. Results for the societal perspective (S9-S11 Figures)
3. References
1. **Model parameter inputs and assumptions**
   1.1. **RSV incidence rate by age and calendar month**

|  | <1 month | 1 month | 2 months | 3 months | 4 months | 5 months | 6 months | 7 months | 8 months | 9 months | 10 months | 11 months |
| --- | --- | --- | --- | --- | --- | --- | --- | --- | --- | --- | --- | --- |
| Sep | 2.46115 | 5.56329 | 5.11227 | 8.03618 | 6.41498 | 7.55968 | 5.37389 | 4.90024 | 6.70796 | 3.34953 | 4.67774 | 5.20172 |
| Oct | 5.4927 | 12.4159 | 11.4094 | 17.9348 | 14.3167 | 16.8714 | 11.9932 | 10.9362 | 14.9706 | 7.47535 | 10.4396 | 11.609 |
| Nov | 7.14052 | 16.1407 | 14.8322 | 23.3153 | 18.6117 | 21.9328 | 15.5912 | 14.217 | 19.4617 | 9.71795 | 13.5715 | 15.0917 |
| Dec | 18.3477 | 41.474 | 38.1116 | 59.9092 | 47.8233 | 56.357 | 40.062 | 36.531 | 50.0074 | 24.9705 | 34.8722 | 38.7785 |
| Jan | 22.6046 | 51.0963 | 46.9539 | 73.8087 | 58.9187 | 69.4323 | 49.3568 | 45.0065 | 61.6096 | 30.7639 | 42.9629 | 47.7755 |
| Feb | 19.2562 | 43.5274 | 39.9986 | 62.8753 | 50.191 | 59.1472 | 42.0455 | 38.3397 | 52.4833 | 26.2068 | 36.5988 | 40.6984 |
| Mar | 15.3796 | 34.7646 | 31.9462 | 50.2175 | 40.0868 | 47.2399 | 33.581 | 30.6213 | 41.9176 | 20.931 | 29.2308 | 32.5052 |
| Apr | 8.23906 | 18.6239 | 17.114 | 26.9022 | 21.4751 | 25.3071 | 17.9898 | 16.4042 | 22.4558 | 11.213 | 15.6594 | 17.4135 |
| May | 3.50688 | 7.92709 | 7.28443 | 11.4507 | 9.14066 | 10.7717 | 7.65722 | 6.98232 | 9.55813 | 4.77272 | 6.66527 | 7.41189 |
| Jun | 1.79569 | 4.05905 | 3.72998 | 5.8633 | 4.68046 | 5.51565 | 3.92086 | 3.57528 | 4.89422 | 2.44386 | 3.41294 | 3.79525 |
| Jul | 0.612648 | 1.38485 | 1.27258 | 2.00042 | 1.59686 | 1.88181 | 1.33771 | 1.2198 | 1.66979 | 0.833789 | 1.16441 | 1.29485 |
| Aug | 0.792217 | 1.79076 | 1.64558 | 2.58675 | 2.06491 | 2.43337 | 1.72979 | 1.57733 | 2.15922 | 1.07817 | 1.50571 | 1.67437 |

**S1 Figure.** RSV incidence rate per 1000 population, by age and calendar month for infants under 1 year born in Ontario, Canada (2009 to 2015).

1.2. **Cost parameters**

**S1 Table.** Cost parameters

| **Parameter** | **Base** | **Range** | **Reference** |
| --- | --- | --- | --- |
| Cost of administration per dose ($) | 17.20 | 0 – 30 | Papenburg *et. al.,*2020; Abu-Raya *et. al.,*2020 (1, 2) |
| **Cost per product dose ($)** | | | |
| RSVpreF | 230 | 50 – 400 | Public listed price, Pfizer |
| Nirsevimab | 750 | 50 – 750 | Public listed price, Sanofi |
| **Cost per patient with RSV managed in inpatient setting per day ($)** | | | |
| Per diem in pediatric general ward | 1538 | 1154 – 1923* | Lanctôt *et. al.,* 2008 (3) |
| Per diem in ICU | 3753 | 2815 – 4691* | Lanctôt *et. al.,* 2008; CIHI, 2016 (3, 4) |
| **Attributable 30-day cost per patient with RSV requiring outpatient healthcare provider visit ($)** | | | |
| <3 months | 253 | 190 – 316* | Rafferty *et. al.,* 2022 (5) |
| 3 - <6 months | 140 | 105 – 175* |  |
| 6m - <1 year | 137 | 103 – 171* |  |
| 1- <2 years | 124 | 93 – 155* |  |
| **Attributable 30-day cost per patient with RSV requiring ED visit ($)** | | | |
| <3 months | 232 | 174 – 290* | Rafferty *et. al.,* 2022 (5) |
| 3 - <6 months | 143 | 107 – 178* |  |
| 6m - <1 year | 136 | 102 – 170* |  |
| 1- <2 years | 130 | 98 – 163* |  |
| **Out-of-pocket costs ($)** | | | |
| Cost of transportation to vaccination | 3.78 | 2.83 – 4.72* | Mitchell *et. al.,* 2017 (6) |
| Cost of transportation to outpatient care | 3.78 | 2.83 – 4.72* |  |
| Cost of transportation to inpatient care | 188 | 14 – 1,986 |  |
| Cost of childcare and home health after inpatient discharge | 339 | 124 – 993 |  |
| Other out-of-pocket costs for patients with RSV managed in inpatient setting (such as over-the-counter medications and other non-transportation or home expenses) | 387 | 37 – 5,404 |  |
| **Caregiver workdays lost** | | | |
| Hospitalization | 7.3 | 1.1 – 68.1 | Mitchell *et. al.,* 2017 (6) |
| Outpatient healthcare provider visit | 2.5 | 0.5 – 5 | Fragaszy *et. al.*, 2018 (7) |
| ED visit | 2.5 | 0.5 – 5 |  |
| Visit healthcare provider for vaccination | 0.5 |  | Assumption |
| **Labour force participation (%)** | | | |
| Age 16+ | Age-specific values |  | Statistics Canada, 2022 (8) |
| Caregiver | 88.6% |  |  |
| **Average employment income ($)** | | | |
| Age 16+ | Age-specific values |  | Statistics Canada, 2022 (9) |
| Caregiver | 62,400 |  |  |

*Range defined as ±25% of the base value

ED: emergency department; ICU: intensive care unit; RSV: respiratory syncytial virus

1.3. **RSV-associated health utility decrements**

**S2 Table.** RSV-associated QALY losses

| **Parameter** | **Base** | **Range** | **Reference** |
| --- | --- | --- | --- |
| **Outpatient primary care visit** | | | |
| Infants | 0.00845 | 0.005 – 0.0454 | Glaser *et. al.,* 2022; Régnier *et. al.,* 2013 (10, 11) |
| Caregivers | 0.00423 | 0.0 – 0.025 |  |
| **ED visit** | | |  |
| Infants | 0.0135 | 0.008 – 0.0454 |  |
| Caregivers | 0.00675 | 0.0 – 0.025 |  |
| **Pediatric general ward hospitalization** | | | |
| Infants | 0.0169 | 0.01 – 0.0726 | Glaser *et. al.,* 2022 (10) |
| Caregivers | 0.0067 | 0.0 – 0.0373 |  |
| **ICU admission** | | | |
| Infants | 0.0245 | 0.0145 – 0.1053 | Roy, 2013 (12); Assumption |
| Caregivers | 0.0097 | 0.0 – 0.0541 |  |

ED: emergency department; ICU: intensive care unit

1.4. **Inputs for higher burden and higher medical costs scenario**

**S3 Table.** Parameters for higher burden and higher medical costs scenario analysis

| **Parameter** | **Base** | **Reference** |
| --- | --- | --- |
| **RSV-associated hospitalization rate** | | |
| Relative hospitalization increase in Northern Canada compared to the rest of Canada | 5 | Karron *et al*., 1999, Singleton *et. al*., 2022 (13, 14) |
| **Average cost per RSV case ($)** |  |  |
| Outpatient visit | 1,747 | Nourbakhsh *et. al.,* 2021(15) |
| Pediatric ward | 18,869 |  |
| ICU | 73,532 |  |
| **Immunization characteristics** | | |
| Cost of administration per dose | 50 | Banerji *et. al.,* 2016; Nourbakhsh *et. al.,* 2021 (15, 16) |
| **Cost of transportation ($)** |  |  |
| All transportation cost (including medical evacuation cost of $16,576) | 18,010 | Banerji et. Al. 2013 (16) |

1.5. **Effectiveness and waning assumptions**

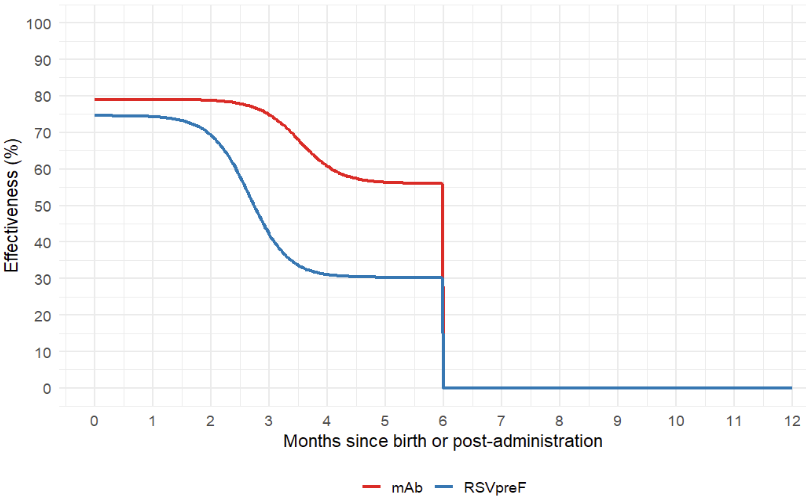

**S2 Figure.** Effectiveness of RSVpreF and monoclonal antibodies (mAb) against Medically Attended Lower Respiratory Tract Infection (MA-LRTI), by month.

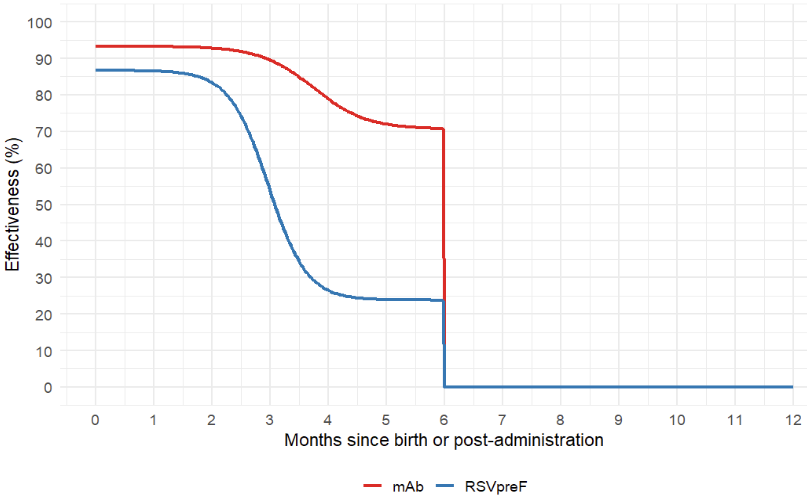

**S3 Figure.** Effectiveness of RSVpreF and monoclonal antibodies (mAb) against hospitalization, by month.

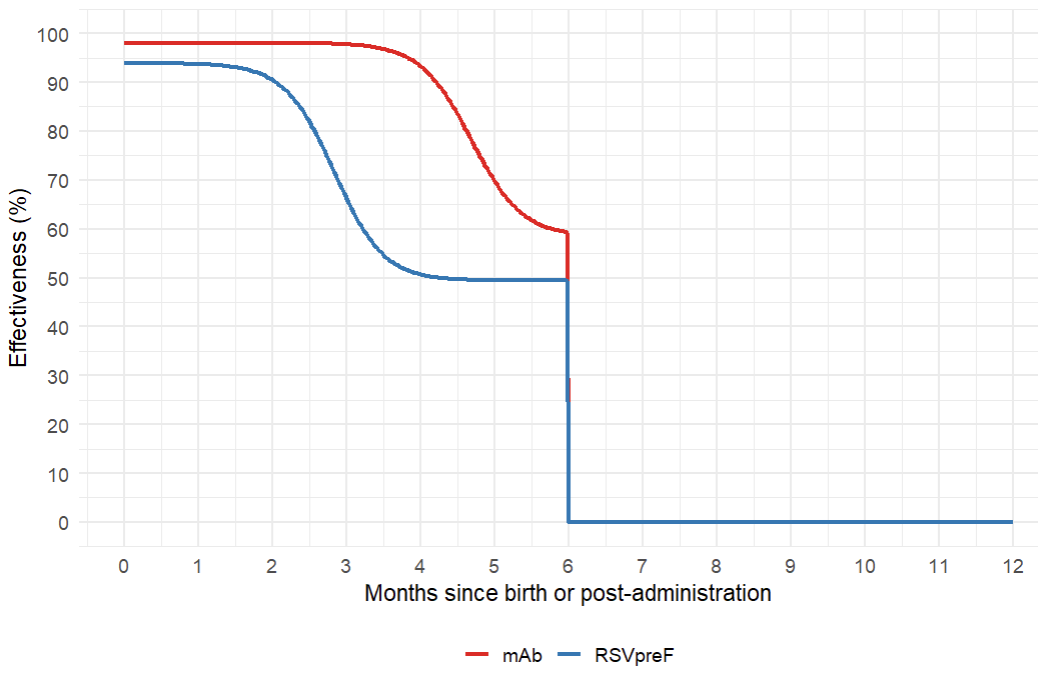

**S4 Figure.** Effectiveness of RSVpreF and mAb effectiveness against intensive care unit admission, by month.

2. **Results**
   2.1. **Primary analysis results**

**S4 Table.** Results of the primary analysis showing costs, QALY losses, and sequential incremental cost-effectiveness ratios (ICERs), in which monoclonal antibody and/or RSVpreF programs are compared stepwise to identify the most efficient options from a health system perspective *

| **Strategies** | **Costs**  **(2024 CAD)** | **QALY loss** | **Sequential ICER** |
| --- | --- | --- | --- |
| Seasonal mAb (moderate or high-risk) + catch-up (moderate or high-risk) | 304,140 | -12.4818 |  |
| Seasonal RSVpreF + seasonal mAb (high-risk) + catch-up (high-risk) | 340,054 | -11.4676 | 35,408 |
| Seasonal RSVpreF at 32-36 weeks of pregnancy for those with in-season deliveries | 378,003 | -14.0409 | dominated |
| Seasonal RSVpreF + seasonal mAb (all unimmunized) + catch-up (high-risk) | 435,265 | -10.7470 | 132,131 |
| Seasonal mAb (all infants) + catch-up (high-risk) | 545,696 | -10.6472 | dominated |
| Seasonal RSVpreF + seasonal mAb (all unimmunized) with catch-up (all infants) | 675,550 | -10.6311 | 1,107,414 |
| Seasonal mAb (all infants) + catch-up (all infants) | 774,523 | -10.5314 | 1,975,226 |

* Monoclonal antibody and/or RSVpreF programs are compared stepwise to identify the most efficient options from a health system perspective All RSVpreF programs are seasonally administered between 32^0/7^ and 36^6/7^ weeks of pregnancy to those with in-season due dates (November to April) i.e., all infants born outside of the RSV season and high-risk infants (born before 32 weeks of gestational age) born during the RSV season were not protected by the RSVpreF pregnancy vaccine. Seasonal monoclonal antibody for “all unimmunized infants” refer to administration of monoclonal antibody (nirsevimab or clesrovimab) both to infants at high-risk and to those not at high-risk who were born to unvaccinated pregnant women and pregnant people during the RSV season. For combination programs, infants received protection from either product (RSVpreF or monoclonal antibody), not both.

2.2. **Secondary analysis results (mAb only programs)**

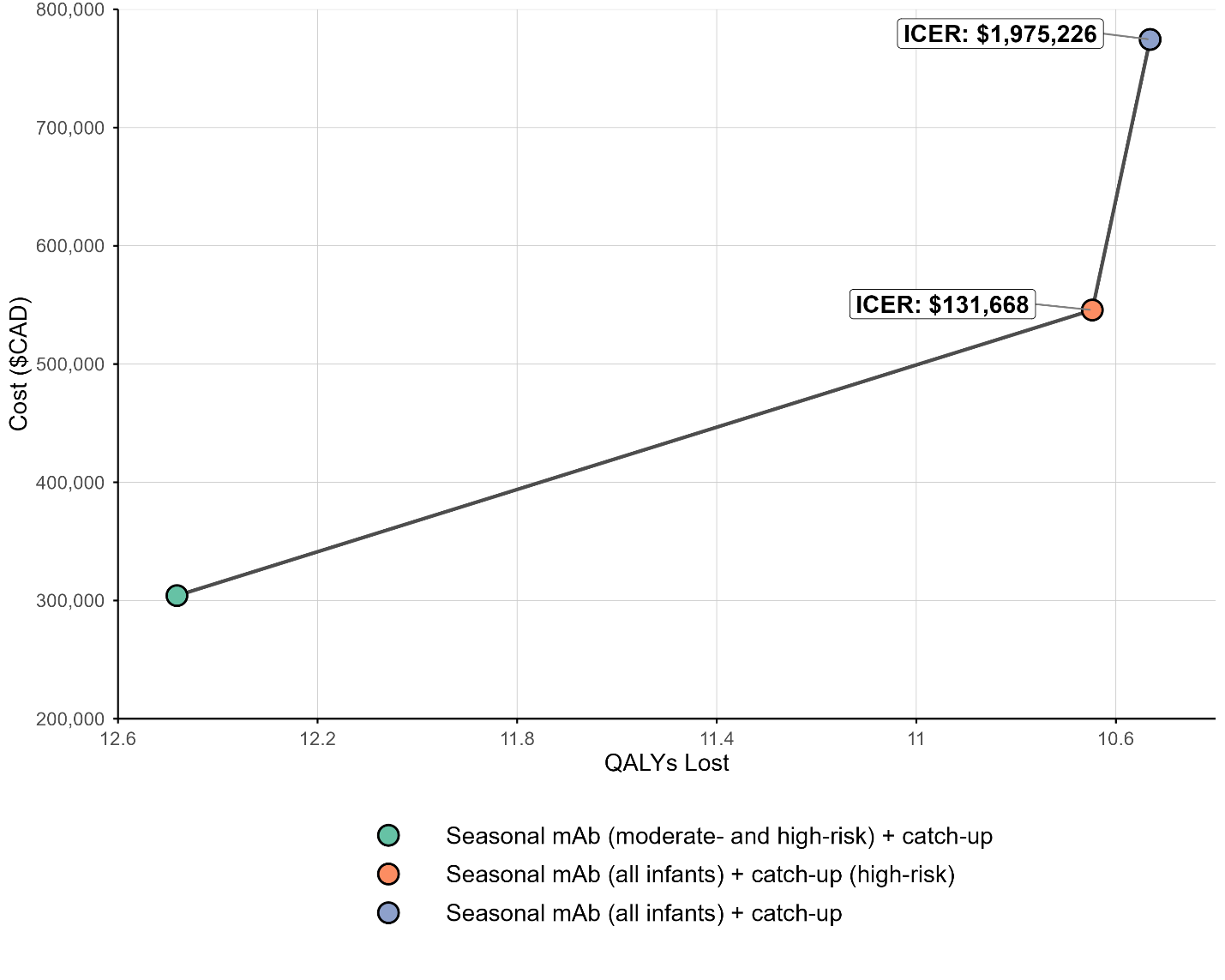

**S5 Figure.** Results of the secondary analysis showing costs, QALY losses, and sequential incremental cost-effectiveness ratios (ICERs), in which monoclonal antibody only programs are compared stepwise to identify the most efficient options from a health system perspective

2.3. **Longer protection duration scenario results**

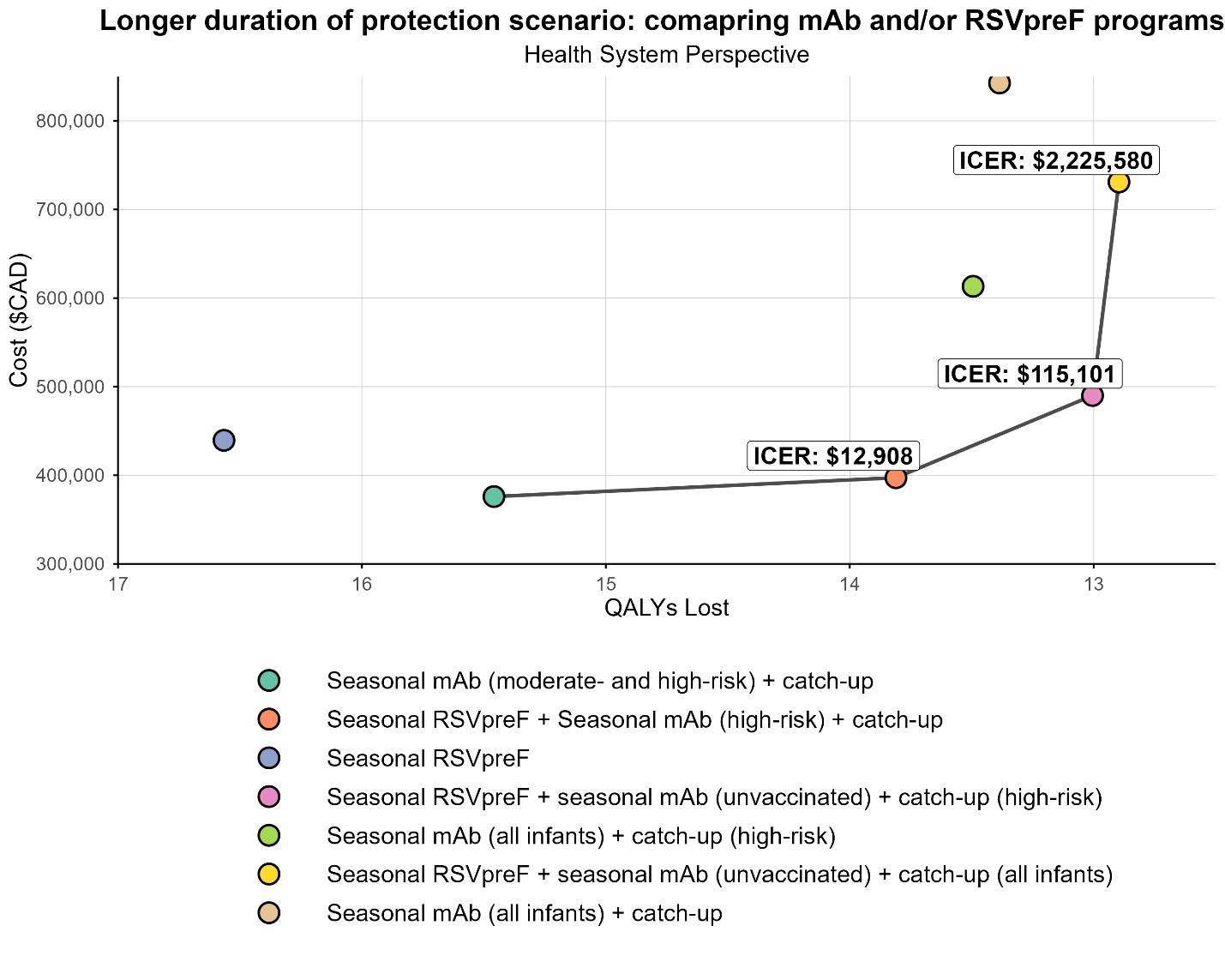

**S6 Figure.** Results of longer duration of protection scenario showing costs, QALY losses, and sequential incremental cost-effectiveness ratios (ICERs) from a health system perspective.

2.4. **Higher RSV burden and higher medical costs scenario results**

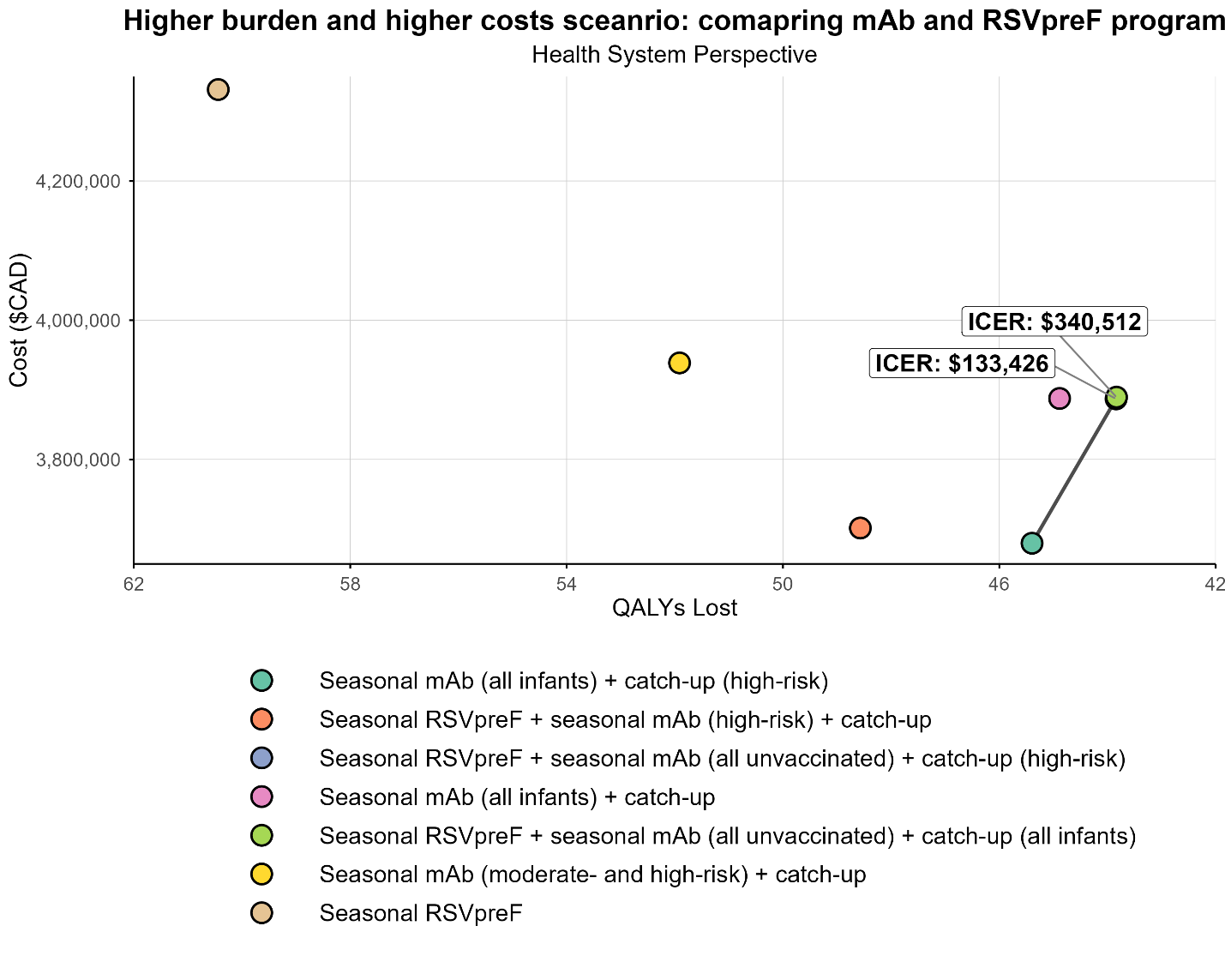

**S7 Figure.** Results of high burden and medical costs scenario showing costs, QALY losses, and sequential incremental cost-effectiveness ratios (ICERs) from a health system perspective. Note: The ICER for the broad combination program (seasonal RSVpreF plus mAb for infants born to unvaccinated pregnant women and pregnant people, with catch-up for high-risk) was $133,426/QALY when compared to broad monoclonal antibodies program (seasonal monoclonal antibodies for all infants with catch-up for infants at high-risk). The ICER for the universal combination program (seasonal RSVpreF plus mAb for all infants born to unvaccinated pregnant women and pregnant people, with catch-up for all infants) was $340,512/QALY, compared to the broad combination program.

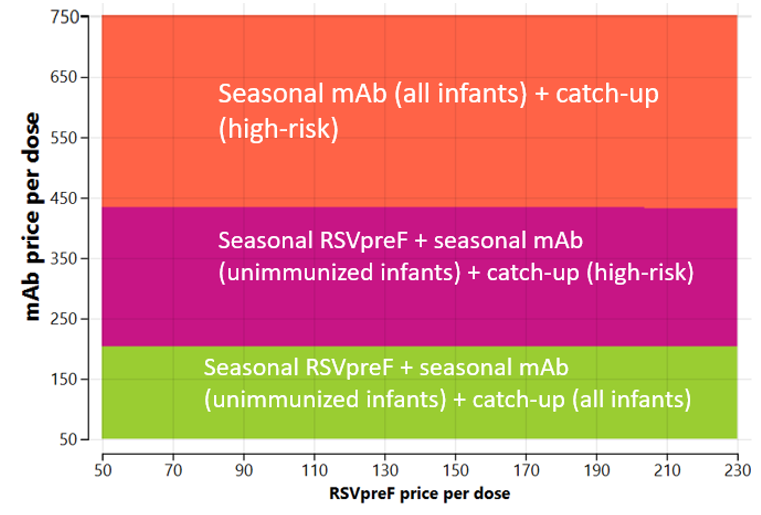

**S8 Figure.** Two-way sensitivity analysis for high burden and higher medical costs setting using a $50,000 per QALY cost-effectiveness threshold from a health system perspective.

2.5. **Results for the societal perspective**

In addition to the health system perspective, a societal perspective was applied to capture the broader economic impact of RSV disease and associated prevention strategies. Under this perspective, additional costs included:

- **Productivity losses due to RSV-related deaths**, calculated using a human capital approach. This method quantified the present value of future earnings lost due to premature mortality among infants, discounted over the expected working lifetime. Age-specific labour force participation rates and average wage data were sourced from Statistics Canada (8, 9).
- **Caregiver productivity losses** associated with time taken off work to care for infants who required medical attention for RSV. Time lost was assumed to vary by healthcare setting (e.g., hospitalization, emergency department, or outpatient primary care visits) and was estimated using average earnings and labour force participation rates for adults aged 25–54 years (8, 9).
- **Out-of-pocket expenses**, including direct non-medical costs borne by families (e.g., over-the-counter medications, home health care expenses, and other miscellaneous household costs related to care) (S1 Table).

These additional costs were incorporated to calculate **total societal costs**, and a separate incremental cost-effectiveness ratio (ICER) was computed for each strategy. complete results from the societal perspective are provided here.

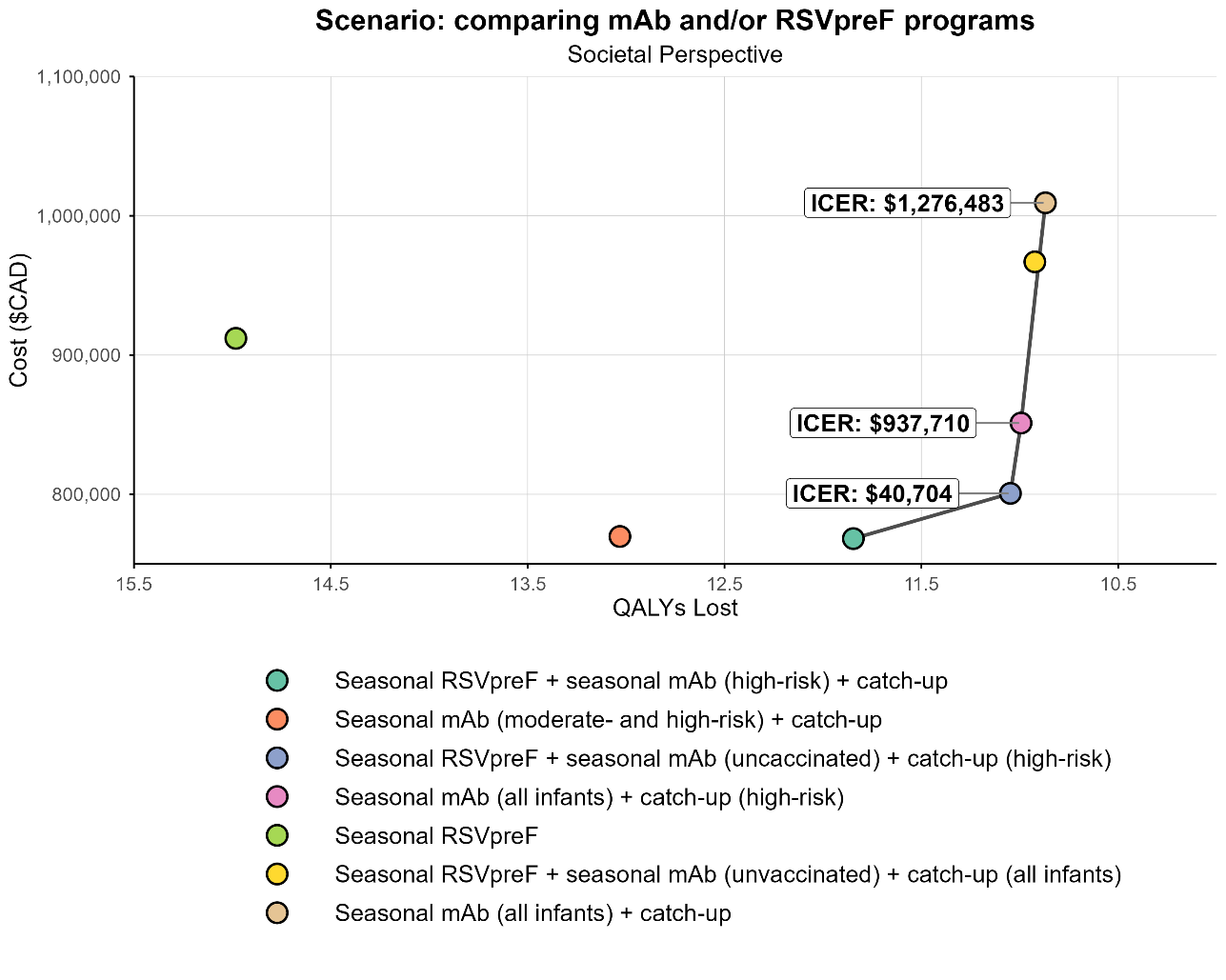

**S9 Figure.** Results of the primary cost-utility analysis showing costs, QALY losses, and sequential incremental cost-effectiveness ratios, in which monoclonal antibody and/or RSVpreF programs are compared stepwise to identify the most efficient options from a societal perspective.

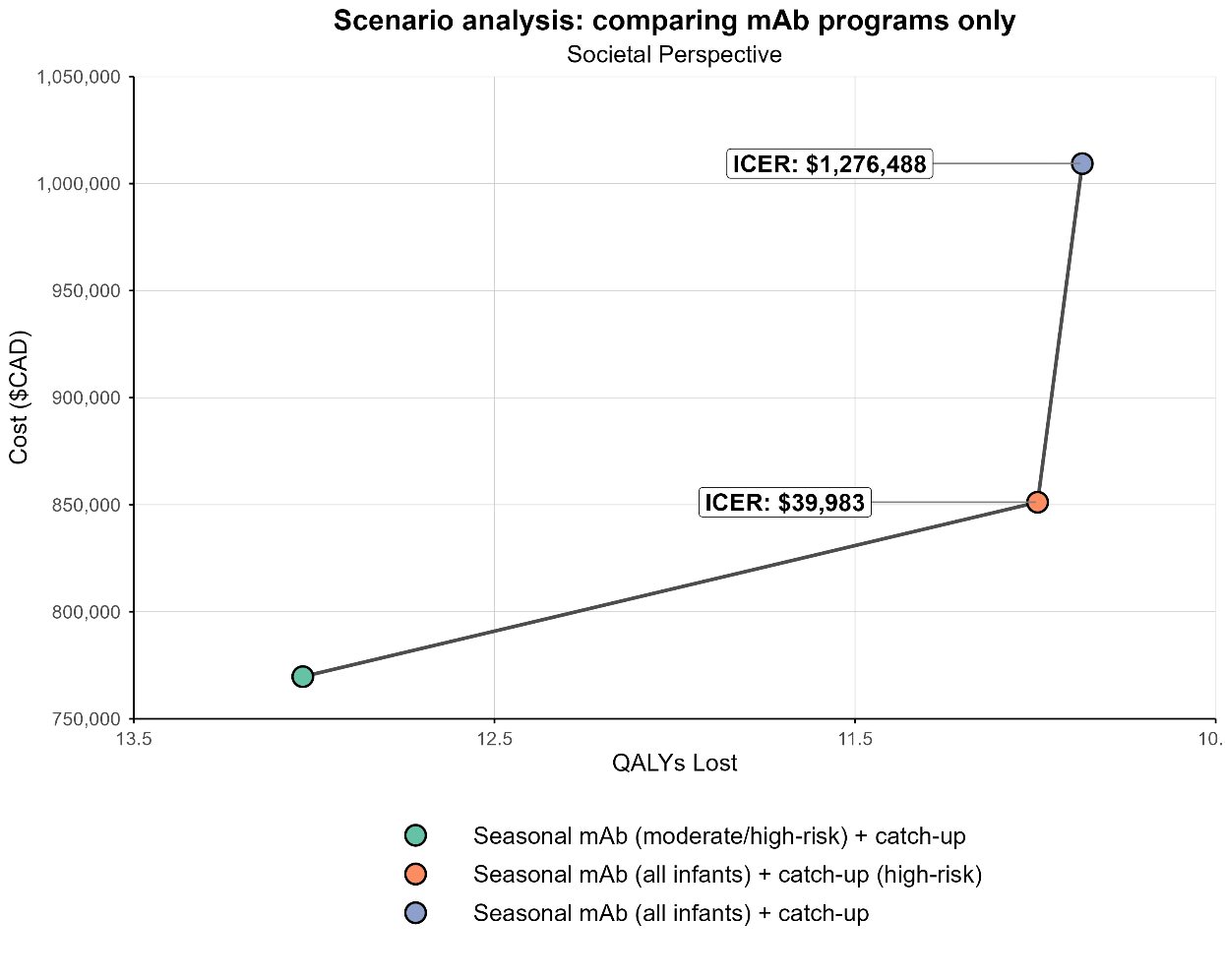

**S10 Figure.** Results of the secondary analysis showing costs, QALY losses, and sequential incremental cost-effectiveness ratios (ICERs), in which monoclonal antibody only programs are compared stepwise to identify the most efficient options from a societal perspective

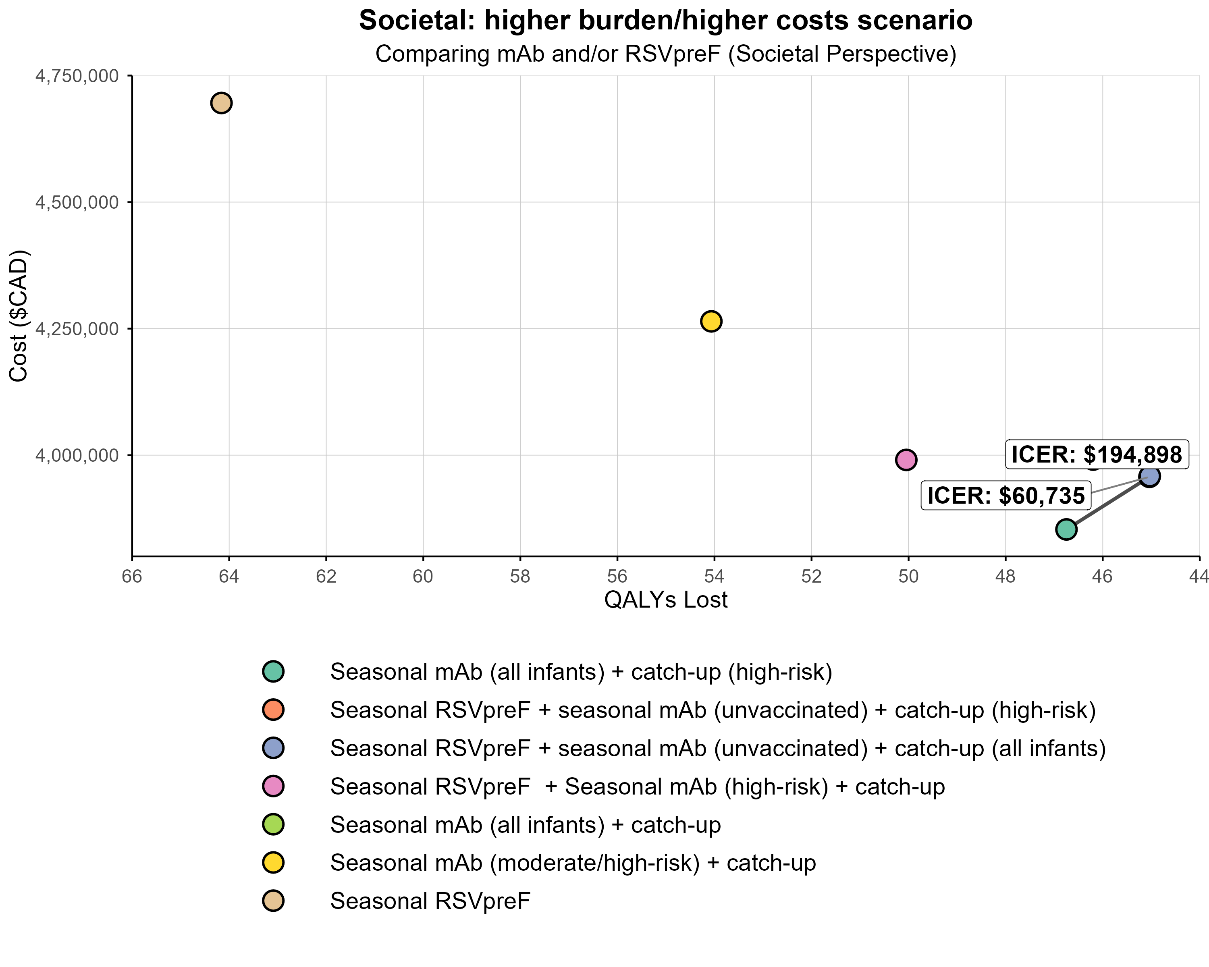

**S11 Figure.** Results of high burden and higher medical costs scenario showing costs, QALY losses, and sequential incremental cost-effectiveness ratios (ICERs) from a societal perspective. Note: The ICER for the broad combination program (seasonal RSVpreF plus mAb for infants born to unvaccinated pregnant women and pregnant people, with catch-up for high-risk) was $60,735/QALY when compared to broad monoclonal antibodies program (seasonal monoclonal antibodies for all infants with catch-up for infants at high-risk). The ICER for the universal combination program (seasonal RSVpreF plus mAb for all infants born to unvaccinated pregnant women and pregnant people, with catch-up for all infants) was $194,898/QALY, compared to the broad combination program.

3. **References**

1.	Abu-Raya B, Coyle D, Bettinger JA, Vaudry W, Halperin SA, Sadarangani M; for members of the Canadian Immunization Monitoring Program, ACTive (IMPACT). Pertussis vaccination in pregnancy in Canada: a cost-utility analysis. CMAJ Open. 2020;8(4):E651-E658. doi: 10.9778/cmajo.20200060.

2.	Papenburg J, Saleem M, Teselink J, Li A, Caouette G, Massé É, Lanctôt KL. Cost-analysis of Withdrawing Immunoprophylaxis for Respiratory Syncytial Virus in Infants Born at 33-35 Weeks Gestational Age in Quebec: A Multicenter Retrospective Study. Pediatr Infect Dis J. 2020;39(8):694-699. doi: 10.1097/INF.0000000000002719.

3.	Lanctôt KL, Masoud ST, Paes BA, Tarride JE, et al. The cost-effectiveness of palivizumab for respiratory syncytial virus prophylaxis in premature infants with a gestational age of 32–35 weeks: a Canadian-based analysis. Curr Med Res Opin. 2008;24(11):3223-37. doi: 10.1185/03007990802484234.

4.	Canadian Institute for Health Information (CIHI). Care in Canadian ICUs. 1–36 (2016).

5.	Rafferty E, Paulden M, Buchan SA, Robinson JL, Bettinger JA, Kumar M, et al. Evaluating the Individual Healthcare Costs and Burden of Disease Associated with RSV Across Age Groups. Pharmacoeconomics. 2022;40(6):633-45.

6.	Mitchell I, Defoy I, Grubb E. Burden of Respiratory Syncytial Virus Hospitalizations in Canada. Can Respir J. 2017;2017:4521302. doi: 10.1155/2017/4521302.

7.	Fragaszy EB, Warren-Gash C, White PJ, Zambon M, Edmunds WJ, Nguyen-Van-Tam JS, Hayward AC; Flu Watch Group. Effects of seasonal and pandemic influenza on health-related quality of life, work and school absence in England: Results from the Flu Watch cohort study. Influenza Other Respir Viruses. 2018;12(1):171-182.

8.	Statistics Canada. Table 14-10-0327-02. Unemployment rate, participate rate and employment rate by sex, annual. DOI: 2022. Available at: <https://doi.org/10.25318/1410032701-eng>.

9.	Statistics Canada. Table 11-10-0239-01  Income of individuals by age group, sex and income source, Canada, provinces and selected census metropolitan areas. DOI: 2022. Available at: <https://doi.org/10.25318/1110023901-eng>.

10.	Glaser EL, Hariharan D, Bowser DM, et al. Impact of Respiratory Syncytial Virus on Child, Caregiver, and Family Quality of Life in the United States: Systematic Literature Review and Analysis The Journal of Infectious Diseases. 2022;226(Suppl 2):S236-S245.

11.	Regnier SA. Respiratory syncytial virus immunization program for the United States: impact of performance determinants of a theoretical vaccine. Vaccine. 2013;31(40):4347-54.

12.	Roy, LM. Deriving health utility weights for infants with Respiratory Syncytial Virus (RSV). University of British Columbia. Thesis/Dissertation, 2013. Accessed 20 June 2023. Available at: <https://open.library.ubc.ca/collections/ubctheses/24/items/1.0074259>.

13.	Singleton R, Semling C, Parker J. Palivizumab Prophylaxis in Alaska for the 2022–23 RSV Season. States of Alaska Epidemiology Bulletin No. 11; 2022. Accessed 25 July 2023. Available at: <https://epi.alaska.gov/bulletins/docs/b2022_11.pdf>.

14.	Karron R SR, Bulkow L et al. Severe respiratory syncytial virus disease in Alaska Native children. J Infect Dis. 1999;180:41–9.

15.	Nourbakhsh S, Shoukat A, Zhang K, Poliquin G, Halperin D, Sheffield H, et al. Effectiveness and cost-effectiveness of RSV infant and maternal immunization programs: A case study of Nunavik, Canada. EClinicalMedicine. 2021;41:101141.

16.	Banerji A, Ng K, Moraes TJ, Panzov V, Robinson J, Lee BE. Cost-effectiveness of palivizumab compared to no prophylaxis in term infants residing in the Canadian Arctic. CMAJ Open. 2016;4(4):E623-E633.
